# Beyond amyloid and tau: synaptic and neurodegenerative biomarkers shape MCI progression

**DOI:** 10.1101/2025.10.06.25337445

**Authors:** Constance Delaby, Susanna Schraen-Maschke, Claire Paquet, Frédéric Blanc, Jean-Sébastien Vidal, Christophe Hirtz, Said Assou, Bernadette Allinquant, Stéphanie Bombois, Audrey Gabelle, Olivier Hanon, Sylvain Lehmann, the Balazar study group

**Affiliations:** LBPC-PPC, Université de Montpellier, INM INSERM, IRMB CHU de Montpellier, Montpellier, France; Univ. Lille, Inserm, CHU Lille, UMR-S-U1172, LiCEND, Lille Neuroscience & Cognition, LabEx DISTALZ, F-59000, Lille, France; Université Paris Cité, UMR-S 1144, APHP, Hospital Broca, Memory Resource and Research Centre of de Paris-Broca-Ile de France, F-75013 Paris, France; Sant Pau Memory Unit, Hospital de la Santa Creu i Sant Pau - Biomedical Research Institute Sant Pau - Universitat Autònoma de Barcelona, Barcelona, Spain; Université de Strasbourg, CHRU de Strasbourg, Memory Resource and Research Centre of Strasbourg/Colmar, French National Centre for Scientific Research (CNRS), ICube Laboratory and Fédération de Médecine Translationnelle de Strasbourg (FMTS), Team Imagerie Multimodale Intégrative en Santé (IMIS)/Neurocrypto, F-67000 Strasbourg, France; Université Paris Cité, GHU APHP Nord Lariboisière Fernand Widal, Centre de Neurologie Cognitive, F-75010 Paris, France; UMR-S1266, Université Paris Cité, Institute of Psychiatry and Neuroscience, Inserm, Paris, France; Assistance Publique-Hôpitaux de Paris (AP-HP), Département de Neurologie, Centre des Maladies Cognitives et Comportementales, GH Pitié-Salpêtrière, Paris, France; Université de Montpellier, Memory Research and Resources center, department of Neurology, Inserm INM NeuroPEPs team, F-34000 Montpellier, France; Institute for Regenerative Medicine and Biotherapy (IRMB), University of Montpellier, INSERM, CHU Montpellier, 34295 Montpellier, France

**Author notes:** **Corresponding author** Pr Sylvain Lehmann, CHU and University of Montpellier, 80 av Fliche, 34295 Montpellier, France.

## Abstract

Accurate prediction of which patients with mild cognitive impairment (MCI) will progress to dementia remains a major challenge. Current biomarkers detect amyloid pathology with high accuracy but offer limited prognostic value for disease progression. We conducted a prospective analysis in the multicentre BALTAZAR cohort, all diagnosed with MCI at baseline and followed for 3 years. Paired cerebrospinal fluid (CSF) and plasma samples were analysed with the NULISA ultrasensitive multiplex platform quantifying more than 120 central nervous system biomarkers. Prognostic performance was assessed using area under the curve (AUC) and hazard ratios (HRs), both for individual markers and for elastic-net-derived biomarker combinations validated by bootstrap and survival analyses. During the 3-year follow-up, 36% of participants converted to dementia. Plasma p-tau biomarkers showed strong accuracy for detecting amyloid positivity (AUC > 0.90) but limited prognostic value for conversion (AUC < 0.75). In CSF, markers of neurodegeneration (tau, NfL) and synaptic dysfunction (NPTX2 encoding the Neuronal Pentraxin 2) predicted conversion with higher accuracy, exceeding p-tau217 performance. The best-performing CSF combination (IL-16, tau, NPTX2) achieved an AUC of 0.86 (95%CI 0.80-0.91) and an HR of 39.8 (95%CI 9.6-165.2). Plasma combinations (p-tau181 or p-tau217 with YWHAG encoding for 14-3-3 protein gamma, a member of the 14-3-3 protein family) provided only modest improvement, likely reflecting the absence of robust synaptic markers in blood. Prognostic assessment of MCI progression to dementia is best achieved through CSF biomarker combinations reflecting neurodegeneration and synaptic dysfunction, complemented by inflammatory markers. These findings emphasize the clinical and pathophysiological relevance of downstream processes beyond amyloid and tau, and support the implementation of multimarker panels for prognosis and therapeutic monitoring.

## Introduction

The recent contribution of biomarkers has greatly advanced the understanding of Alzheimer’s disease (AD) and has led to its definition as a clinico-biological continuum ^1^. A key discovery within this framework is the ability to detect disease-specific markers more than a decade before the onset of clinical symptoms ^2^. It is noteworthy that this conceptual evolution could already have been anticipated based on neuropathological evidence ^3, 4^. Incidentally, the detection in the cerebrospinal fluid (CSF) of β-amyloid peptide (Aβ42) or its ratio with Aβ40, together with total tau and phosphorylated tau at threonine 181 (p-tau181), has become a major element in the diagnosis of AD. These biomarkers have been integrated into international diagnostic criteria for the disease ^1, 5^ and are now routinely interpreted for clinical reporting of the diagnosis to practitioners ^6^.

Changes in these fluid biomarkers have been associated with the AD pathophysiological AT(N) classification ^7^. The amyloid status (A) is clearly linked to changes in CSF Aβ, which correlate well with amyloid PET imaging. Total tau is strongly associated with the neurodegeneration status (N), as are other biomarkers such as neurofilament light chain (NfL), which can also be elevated in other disorders, including Creutzfeldt–Jakob disease ^8^. For the AD-related tauopathy status (T) in AD, defined histopathologically and by tau PET, CSF p-tau181 is not a perfect surrogate, as it also correlates with neurodegeneration (N) and shows strong correlation with total tau ^9^.

The possibility to detect these biomarkers in blood with strong analytical performance has greatly increased interest in their use for AD. This advance has also enabled refinement of the temporal sequence within the AT(N) framework, with amyloid pathology (A) identified as the earliest detectable event. Tau pathology (T) has also been shown to emerge very early, whereas neurodegeneration (N) occurs later, likely associated with the formation and burden of neurofibrillary tangles. The association of fluid biomarkers with the AT(N) status differs somewhat between blood and CSF. While Aβ remains a specific indicator of amyloid pathology, its performance is reduced due to challenges in accurate measurement in this fluid. Moreover, peripheral production of Aβ may dilute the brain-derived signal, further reducing its diagnostic precision. Neurodegenerative biomarkers such as tau and NfL face similar limitations, as their levels can be influenced by production outside the brain and by non-AD conditions ^10, 11^. Encouraging results came from p-tau181, and even more from p-tau217, which initially demonstrated superior diagnostic performance for AD in CSF ^12^. In blood, p-tau217 has emerged to be an early and highly sensitive marker of amyloid status ^13–16^. Its close association to amyloid pathology was further confirmed in therapeutic trials, where p-tau217 proved to be a robust surrogate marker of treatment impact ^17, 18^.

However, in predicting the conversion of mild cognitive impairment (MCI) to dementia stage, the performance of current biomarkers remains modest ^19, 20^, as previously reported in the BALTAZAR cohort ^13, 21^. To address this limitation, we applied a targeted proteomic approach using NULISA technology. Using its CNS panel, NULISA enables the simultaneous exploration of biomarkers related not only to amyloid and tau pathology, but also to neurodegeneration, synaptic dysfunction, neuroinflammation, and immune responses ^22^. This technology has recently shown strong biomarkers detection capabilities across different cohorts, though these studies were not designed, as BALTAZAR was, to investigate MCI prognosis ^22–24^. Moreover, previous NULISA investigations were restricted to blood analyses. By assessing biomarkers in both CSF and plasma, our objective was to identify those most informative for predicting conversion. In this study, we emphasize the central role of neurodegeneration together with synaptic impairment and neuroinflammation, in driving conversion. We also propose biomarker combinations with improved prognostic performance.

## Methods

### BALTAZAR study population

The study population consisted of 173 participants from the BALTAZAR multicenter prospective cohort (ClinicalTrials.gov Identifier: NCT01315639) ^25^ who underwent lumbar puncture as part of the protocol. All participants underwent clinical, neuropsychological, imaging, and biological assessments. APOE was genotyped in a single centralized laboratory. Mild cognitive impairment (MCI) subjects were diagnosed according to Petersen’s criteria ^26, 27^. Participants were followed every six months for three years, with reassessments conducted at each visit to monitor cognitive decline and progression to a dementia stage ^25^. The conversions from MCI to dementia were reviewed by an adjudication committee based solely on clinical and neuropsychological characteristics, using the National Institute on Aging–Alzheimer’s Association (NIA-AA) criteria, with assessors blinded to CSF and plasma biomarker results. Progression from MCI to dementia was defined by a decline in cognitive functioning and increased disability in activities of daily living, as measured by changes from baseline in scores on the Mini-Mental State Examination (MMSE), Instrumental Activities of Daily Living (IADL), Activities of Daily Living (ADL), and the Clinical Dementia Rating (CDR ≥ 1) scale ^27^.

### ELISA, Quanterix and Lumipulse measurement in CSF and plasma

CSF and plasma samples were collected at the first visit, and to minimize pre-analytical and analytical variability, identical collection tubes were used across all centers. Aliquots were stored at –80 °C in low-binding Eppendorf® LoBind microtubes (Eppendorf, ref. 022431064, Hamburg, Germany) until testing.

Biomarker levels of CSF tau, p-tau181, Aβ40, and Aβ42 were measured using standardized, commercially available EU-approved *in vitro* diagnostic (IVD) ELISA kits with performance comparable to reference assay ^28^ (Euroimmun β-amyloid 1–40 and 1–42 ^29^, Innotest hTau, and Innotest Phospho-Tau (181P) ^30^). CSF p-tau217 was determined using the commercial MSD (Meso Scale Discovery, Rockville, MD, USA) S-PLEX Human Tau (pT217) Kit. CSF BACE1 and neurogranin (NRGN) were measured using ELISAs commercialized by Euroimmun, which include ready-to-use lyophilized calibrators and a standardized protocol, as previously reported ^9^.

Plasma Nfl, p-tau181 and p-tau217 were also determined, using ultrasensitive Simoa technology, on an Quanterix HD-X analytical platform (Quanterix™, Billerica, MA, USA). Plasma pTau181 and Nfl with a commercial Advantage V1 kit and pTau217 with the ALZpath assay.

### NULISA measurements

CSF and plasma samples were thawed and centrifuged for 10 min at 10,000 g to remove residuals before transferring 25 µL onto the assay plate (NULISA CNS panel, ref. 8000104). Reagent plates were homogenized with 15 upside-down rotations. Sample, library, and reagent plates were then centrifuged for 20 sec at 352 g prior to loading onto the ARGO HT System (ref. 8000101). After the run, the prepared NULISA sequencing library tube was stored at –20 °C until sequencing. Three microliters (3 µL) of NULISA sequencing libraries were used to determine DNA concentrations with the Qubit™ dsDNA HS Assay Kit (Invitrogen by Thermo Fisher Scientific, USA) on a QFX spectrofluorometer (DeNovix Inc.™, USA). Libraries were then sequenced using a NextSeq 500/550 High Output Kit v2.5 (20024906, Illumina) on an Illumina NextSeq 550 System at the Transcriptome Core Facility of the Institute for Regenerative Medicine and Biotherapy, CHU–Inserm–UM Montpellier, France.

Output files were processed by the Alamar cloud system to generate NULISA protein quantification (NPQ) raw data. Original NPQ values were first linearized using the formula 2(NPQ–1), then divided by the initial protein concentration. The resulting ratio was incremented by 1 to avoid log transformation of zero or negative values, and the result was log2-transformed. Samples were randomized across assay plates, each of which included quality controls. Limits of detection (LOD) were defined as the mean + 3 SD of unlogged normalized counts from negative controls, then rescaled and log2-transformed. Assay reproducibility was assessed by within- and between-run coefficients of variation (CVs) based on sample control replicates.

For consistency and ease of cross-referencing, all biomarkers were reported using uppercase, non-italicized gene symbols, as provided by the manufacturer (*Supplementary Figure 1*). CSF and plasma detectability were defined as the percentage of values below the LOD. We excluded from analysis proteins with detection below 40% (*<u>Supplementary File 1</u>*). For APOE4, detectability was computed only in E4-positive patients identified by genotyping ^25^. To calculate the Aβ42/Aβ40 ratio from NULISA data (Ab4240), the log2 NPQ value of Aβ42 was subtracted from that of Aβ40.

### Statistical analyses

In the BALTAZAR MCI cohort, population characteristics across amyloid status and conversion outcomes were described using percentages (%) for categorical variables and means with standard deviations for continuous variables (M ± SD). Following assessment of variance homogeneity, continuous variables were compared using Student’s or Welch’s t-test as appropriate, and categorical variables using the χ² test.

All NULISA biomarker concentrations were log2-transformed, and biomarkers with more than 40% missing values were excluded. The remaining missing values were imputed using the median. No additional normalization was applied, as standardization was handled by the statistical models.

To compare NULISA data in the MCI BALTAZAR population across amyloid statuses and conversion outcomes, we used linear mixed models adjusted for age, sex, and APOE ε4 carrier status. The false discovery rate (FDR) was controlled using the Benjamini–Hochberg method.

To combine NULISA biomarkers, we first computed individual AUC values using 100 bootstrap iterations, each involving resampling with replacement and fitting a penalized logistic model (alpha = 0.5) with 5-fold cross-validation. The top 20 most frequently selected biomarkers were retained and analyzed by logistic regression with elastic net regularization, combining two or three of them through repeated bootstrap estimation (R = 30) of AUC to assess their predictive performance. The ten best-performing biomarkers and combinations were then re-evaluated using 300 bootstrap replicates to estimate the AUC with 95% confidence intervals, along with model coefficients. They were also used as tertiles in a Kaplan–Meier survival analysis calculated with the log-rank test. Hazard ratios with 95% confidence intervals and corresponding statistical tests were then computed between the 1st and 3rd tertiles.

All analyses were performed using MedCalc (v20.118) and R software (R Core Team, 2019).

## Results

### Demographics of the BALTAZAR population

The BALTAZAR cohort is composed of MCI patients, of whom 38.7% were men, 38.2% carried one or two APOE4 alleles, their mean educational level was 5.5 years (SD = 1.5), and their mean age was 77.8 years (SD = 5.5). Amyloid-positive (Aβ+) participants, defined by the CSF Aβ42/40 ratio ^31^), represented 55.5% of the population. Aβ+ participants did not differ from Aβ–participants in age, sex, hippocampal volume, or educational level (*Table 1*). However, they were more likely to carry the APOE4 allele (56.3% vs 15.6%; P = 0.0036), had a lower MMSE score (26.1 vs 27.1; P = 0.0109), and showed a faster cognitive decline (–1.6 (3.3) vs –1.0 (2.8) MMSE/year; P = 0.0070). Overall, 36% of the MCI population converted to dementia stage during the three-year follow-up. The conversion rate was 50% in the Aβ+ population and 19% in the Aβ− population. MCI converters did not differ from non-converters in age, sex, or educational level (*Table 1).* However, they were more likely to carry the APOE4 allele (52.4% vs 30.0%; P < 0.0001), had a lower hippocampal volume (4.1 (SD 1.1) cm³ vs 5.0 (SD 1.1); P < 0.0001), a lower MMSE score (25.6 (2.5) vs 27.1 (2.2); P = 0.0001), and a faster cognitive decline (–2.9 (4.3) vs –0.4 (1.5) MMSE/year; P < 0.0001).

**Table 1.** Characteristics of the MCI population and subgroup comparisons according to amyloid status and MCI conversion to dementia within 3 years.

| | n | Value | n<br>Amyloid<br>negative | Value | n<br>Amyloid<br>positive | Value | Positive vs<br>Negative (p<br>or $\chi^2$) | n Non<br>Converter | Value | n MCI<br>Converter | Value | Converters vs<br>non<br>converters (p<br>or $\chi^2$) |
| --- | --- | --- | --- | --- | --- | --- | --- | --- | --- | --- | --- | --- |
| Age (years) | 173 | 77.8 (5.5) | 77 | 76.9 (4.8) | 96 | 78.5 (5.8) | 0.0672 | 110 | 77.5 (5.1) | 63 | 78.2 (6) | 0.4039 |
| Men (%) | 173 | 38.7 | 77 | 42.9 | 96 | 35.4 | 0.3195 | 110 | 37.3 | 63 | 41.3 | 0.6046 |
| 1 or 2 APOE4 alleles (%) | 173 | 38.2 | 77 | 15.6 | 96 | 56.3 | <0.0001 | 110 | 30.0 | 63 | 52.4 | 0.0036 |
| Educational level (years) | 171 | 5.5 (1.5) | 76 | 5.4 (1.5) | 95 | 5.6 (1.5) | 0.4972 | 108 | 5.6 (1.5) | 63 | 5.3 (1.6) | 0.3201 |
| Hippocampal volume (R+L) (cm<sup>3</sup>) | 148 | 4.7 (1.1) | 66 | 4.7 (1.3) | 82 | 4.6 (1) | 0.3791 | 93 | 5.0 (1.0) | 55 | 4.1 (1.1) | <0.0001 |
| MMSE (/30) | 170 | 26.5 (2.4) | 76 | 27.1 (2.1) | 94 | 26.1 (2.6) | 0.0109 | 107 | 27.1 (2.2) | 63 | 25.6 (2.5) | 0.0001 |
| MMSE / year | 172 | -1.33 (3.07) | 77 | -1 (2.8) | 95 | -1.6 (3.3) | 0.0070 | 109 | -0.4 (1.5) | 63 | -2.9 (4.3) | <0.0001 |

Legend: Data are presented as mean (SD) for continuous variables and percentages for categorical variables. n indicated the number in each group. Comparisons between amyloid-positive and amyloid-negative groups, as well as between converters and non-converters, were performed using Student's t test or Chi-square ( $\chi^2$ ) test, as appropriate.
Abbreviations: MCI, Mild Cognitive Impairment; APOE4, Apolipoprotein E ε4 allele; R+L, right plus left hippocampal volume; cm<sup>3</sup>, cubic centimeters, MMSE, mini-mental state examination.

### Targeted NULISA proteomics measurement in CSF and plasma, detectability and comparison with other analytical methods

The CNS NULISA panel is composed of biomarkers already identified as relevant to neurodegenerative diseases and spanning multiple pathological processes. The complete list of the panel is provided in *Supplementary Table 1* and are distributed as follows: 52 inflammatory biomarkers, 24 specific to neurodegeneration, 12 related to vascular and metabolic processes, 21 associated with synuclein and synaptic function, and 22 linked to amyloid and tau pathology. Two CSF samples were not properly detected on the NULISA plate and were therefore excluded from the study. The following ten biomarkers were detected in less than 40% of the CSF samples and were therefore not included in the analysis: neprilysin (MME), phosphoglycerate kinase 1 (PGK1), ficolin-2 (FCN2), serum amyloid A-1 protein (SAA1), brain-derived neurotrophic factor (BDNF), protein S100-A12 (S100A12), C-C motif chemokine 17 (TARC/CCL17), CD40 ligand (CD40L/TNFSF5), hemoglobin subunit alpha 1-2 (HBα1-2), and periostin (POSTN). Only one biomarker, pleiotrophin (PTN), was likewise excluded from the plasma analysis.

Biomarker CSF levels of Aβ40, Aβ42, tau, p-tau181, p-tau217, neurogranin, and BACE1 were previously determined in BALTAZAR with alternative assays ^9^. This allowed us to test how these values correlated with those obtained through NULISA (*Figure 1*). All of them showed a significant Spearman correlation (P < 0.001), with rho > 0.8 for Aβ42, Aβ42/40, tau, p-tau181, and p-tau217, the lowest being BACE1 with rho = 0.52. At the lowest concentrations, the flattening of the curves, in particular for Aβ42 and p-tau217, suggests a lower limit of detection for NULISA.

**Figure 1:**
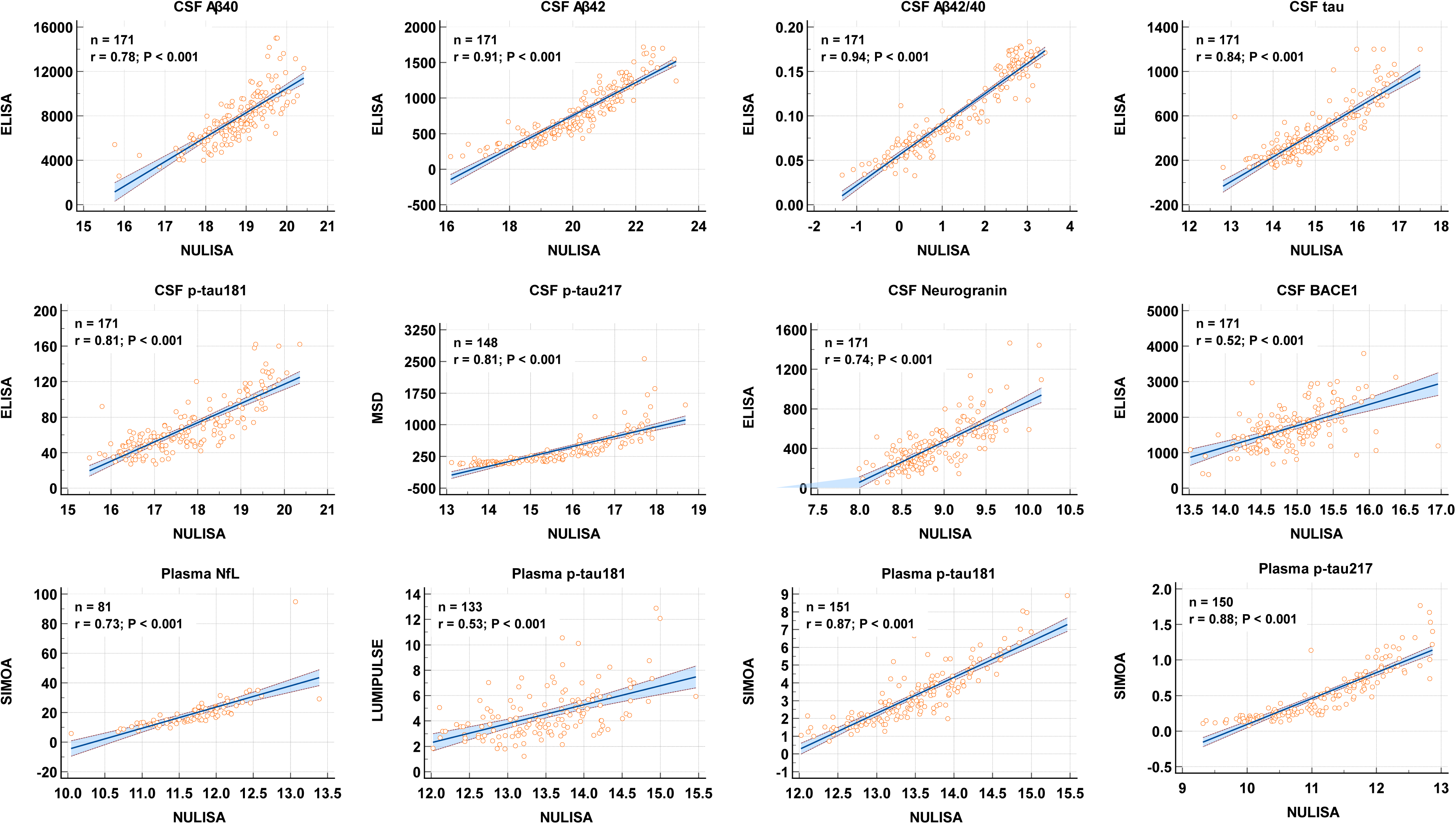
Comparison of NULISA with ELISA, SIMOA, MSD and LUMIPULSE methods. Correlation plots between NULISA and reference methods (ELISA, SIMOA, Lumipulse) for CSF and plasma biomarkers. The number of samples used for each plot is indicated (n), together with the Spearman rank correlation coefficient (r) and corresponding p-value. Strong agreement was observed for Aβ42, Aβ42/40, total tau, p-tau181, and p-tau217, supporting the analytical validity of the NULISA platform. NULISA concentrations are expressed on a log2 scale. Correlation results for all NULISA biomarkers are provided in Supplementary Table 2. Abbreviations: CSF, cerebrospinal fluid; NULISA, Nucleic acid–Linked Immuno-Sandwich Assay; SIMOA, Single molecule array.

In plasma, NfL, p-tau181, and p-tau217 were also previously measured with alternative methods ^13, 21, 32^, and their comparison with NULISA revealed significant correlations across all biomarkers. This was most pronounced for p-tau217 (rho = 0.88), with the curve suggesting that NULISA has a lower detection limit than SIMOA.

As the NULISA panel was assessed in paired CSF and plasma samples, it was possible to correlate the values in both fluids (*Figure 2* and *Supplementary Table 2*). Importantly, all tau-related biomarkers were significantly correlated, the highest being p-tau217 (rho = 0.79; P < 0.001) and the lowest MAPT (corresponding to total tau) (rho = 0.407; P < 0.001). Other neurodegenerative and inflammatory biomarkers such as NEFL, GFAP, CHIT1, and GDF15 were significantly correlated, while the synaptic biomarkers NRGN, NPTX2, and BACE1 were not. Among the biomarkers, CRP showed the strongest correlation between CSF and plasma (rho = 0.86; P < 0.001).

**Figure 2:**
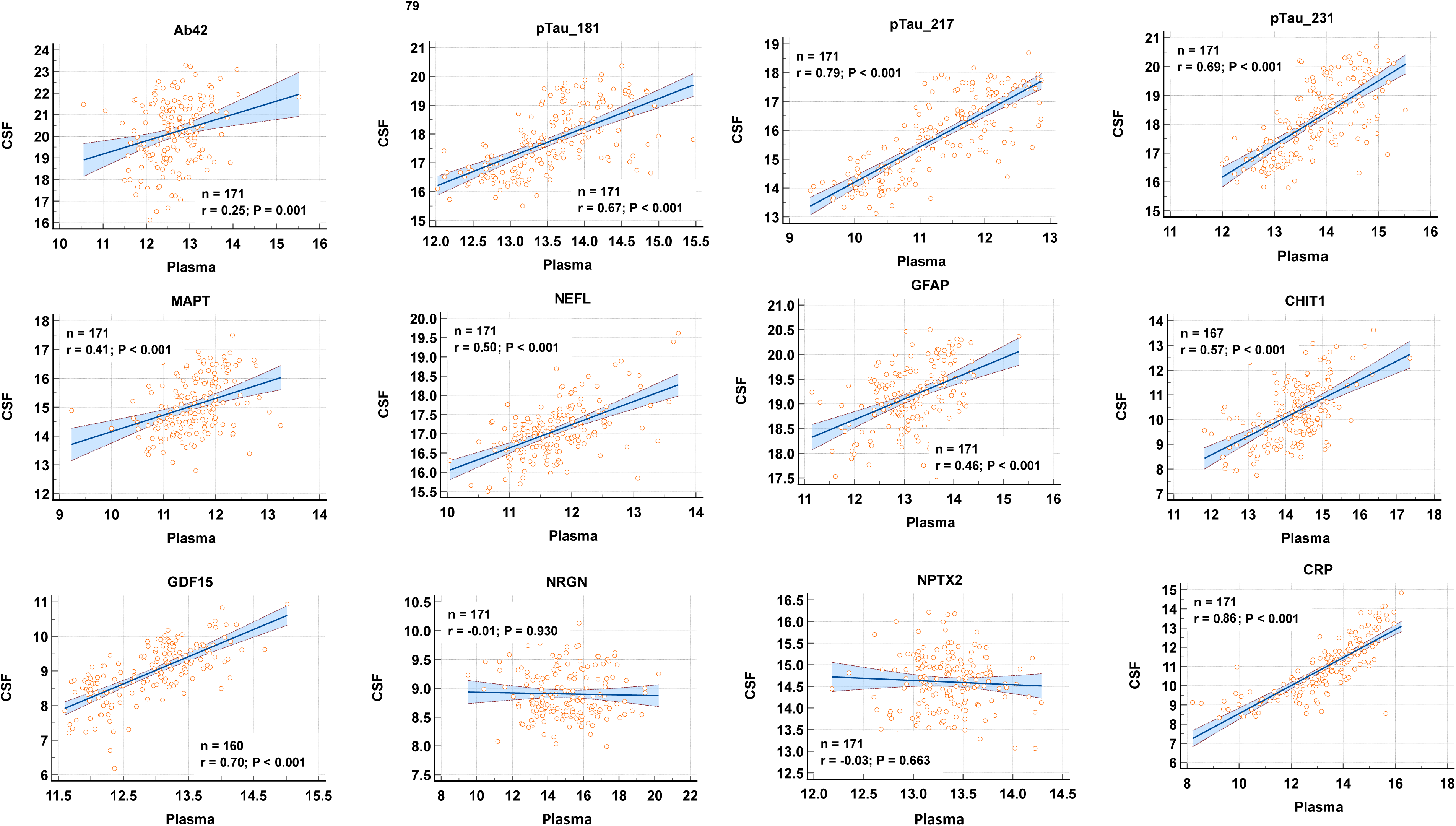
Correlation of NULISA CSF and plasma biomarker concentrations. Scatterplots of matched CSF and plasma measurements across the BALTAZAR cohort. The number of samples used for each plot is indicated (n), together with the Spearman rank correlation coefficient (r) and corresponding p-value. CRP, p-tau biomarkers, NfL (NEFL), and GFAP showed significant correlations between fluids, while synaptic markers such as NPTX2 and neurogranin (NRGN) did not. Correlation data on all the NULISA biomarkers are provided in the Supplementary Table 2.

### Individual CSF and plasma biomarkers in amyloid positive (Aβ+) population

As illustrated in the volcano plots of *Figure 3A* and detailed in *Supplementary Table 3*, CSF tau-related biomarkers were increased in Aβ+, with the following decreasing performance: p-tau217 > p-tau231 > p-tau181 > MAPT. Aβ42 and the Aβ42/40 ratio in CSF also showed high performance but were much lower in plasma, where only the Aβ42/40 ratio remained significant (*Figure 3B*). GFAP was also differential and showed higher performance in plasma than in CSF. In the CSF, other biomarkers had significant p-values, even after adjustment for ApoE, sex, and age, and for multiple comparisons. Several were linked to neurodegeneration, such as NEFL, S100B, or UCHL1. Others were synaptic biomarkers such as SNCA, SNCB, and NRGN, or were linked to glial/microglial cells, neuroinflammatory processes, and metabolic processes, such as MDH1, VSNL1, GFAP, CHIT1, FABP3, and SMOC1. Boxplots illustrating the distribution between Aβ+ and Aβ– are shown in *Figure 4*, upper part.

**Figure 3:**
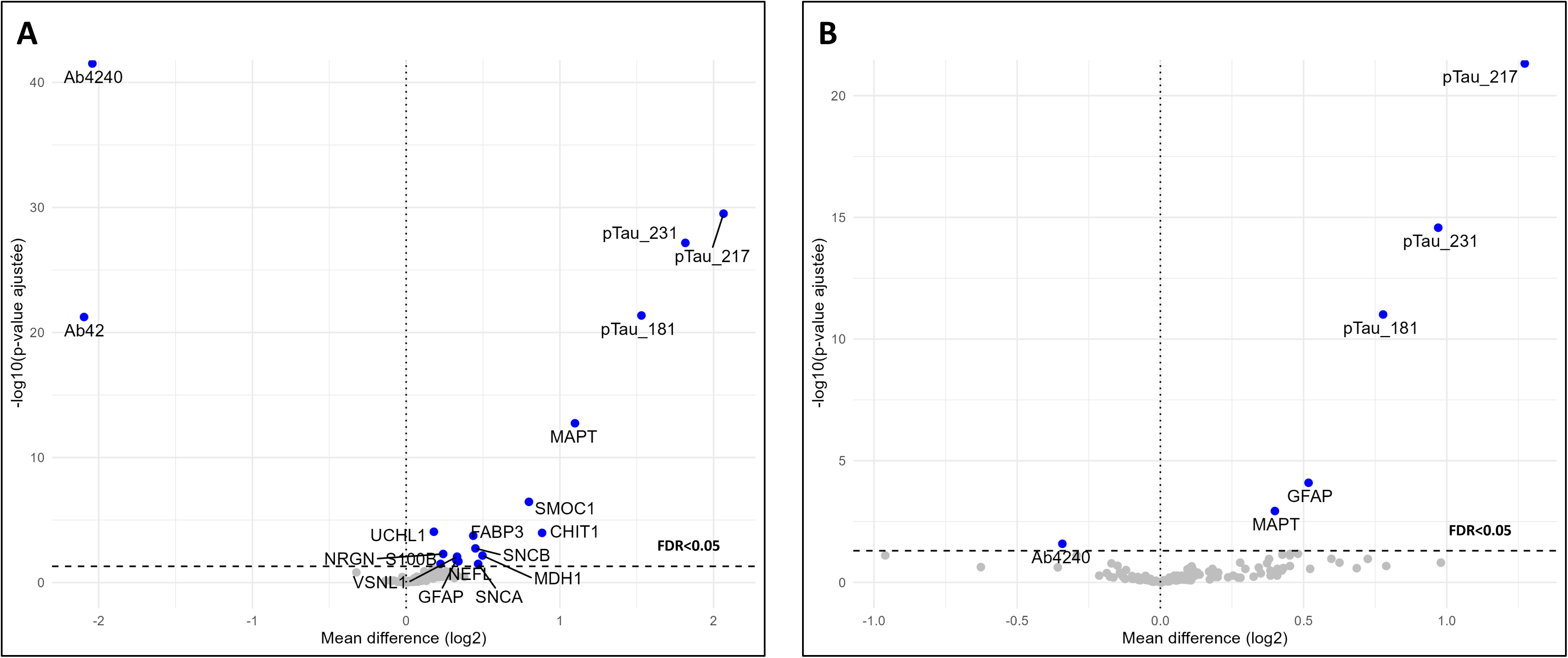
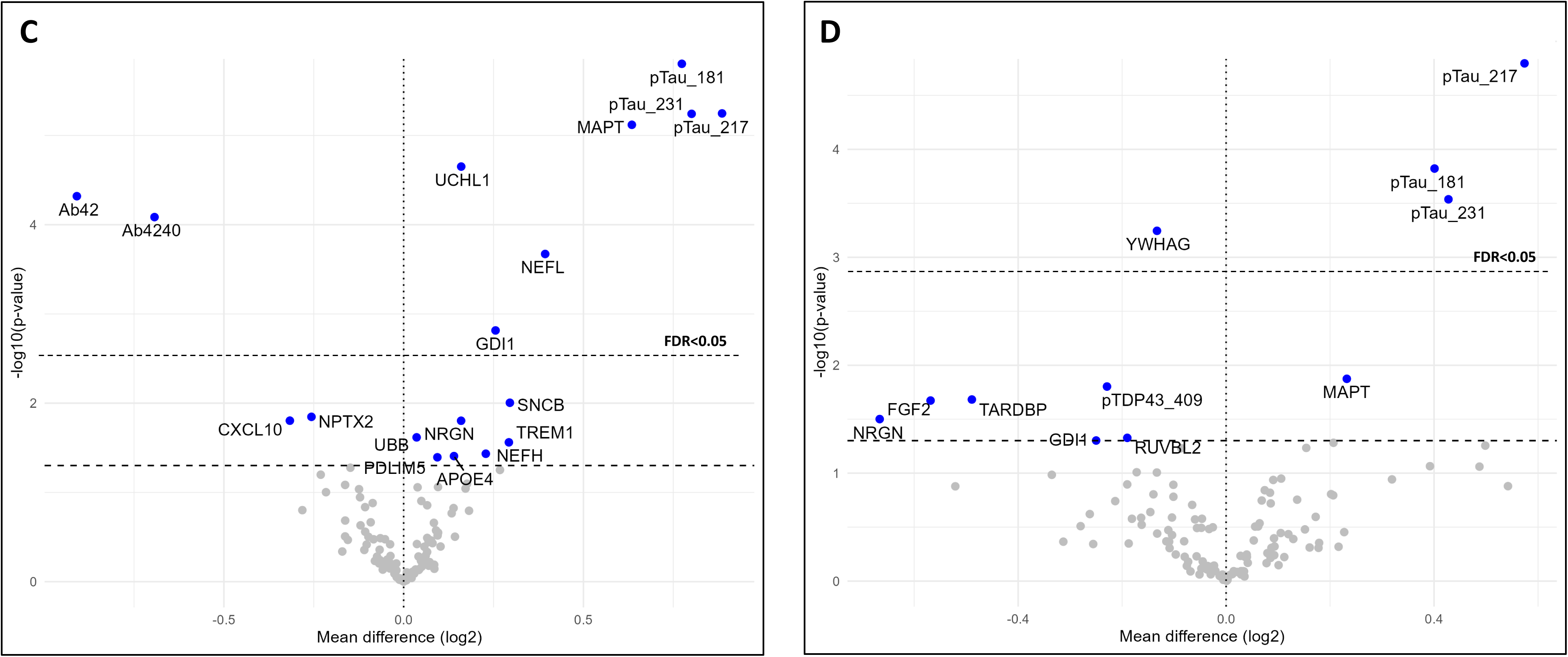
Individual biomarkers associated with amyloid status and MCI conversion. Volcano plots −log10(p-value) versus log2(fold change) illustrating the association between CSF (A, C) and plasma (B, D) NULISA biomarkers with Aβ+ (A, B), and MCI conversion (C, D). In C, D, the limit for false discovery rate (FDR) of 0.05 is indicated. p-values are derived from using adjusted linear mixed models adjusted for age, sex, and APOE4 genotype. The numeric values are reported in the supplementary table 3.

**Figure 4:**
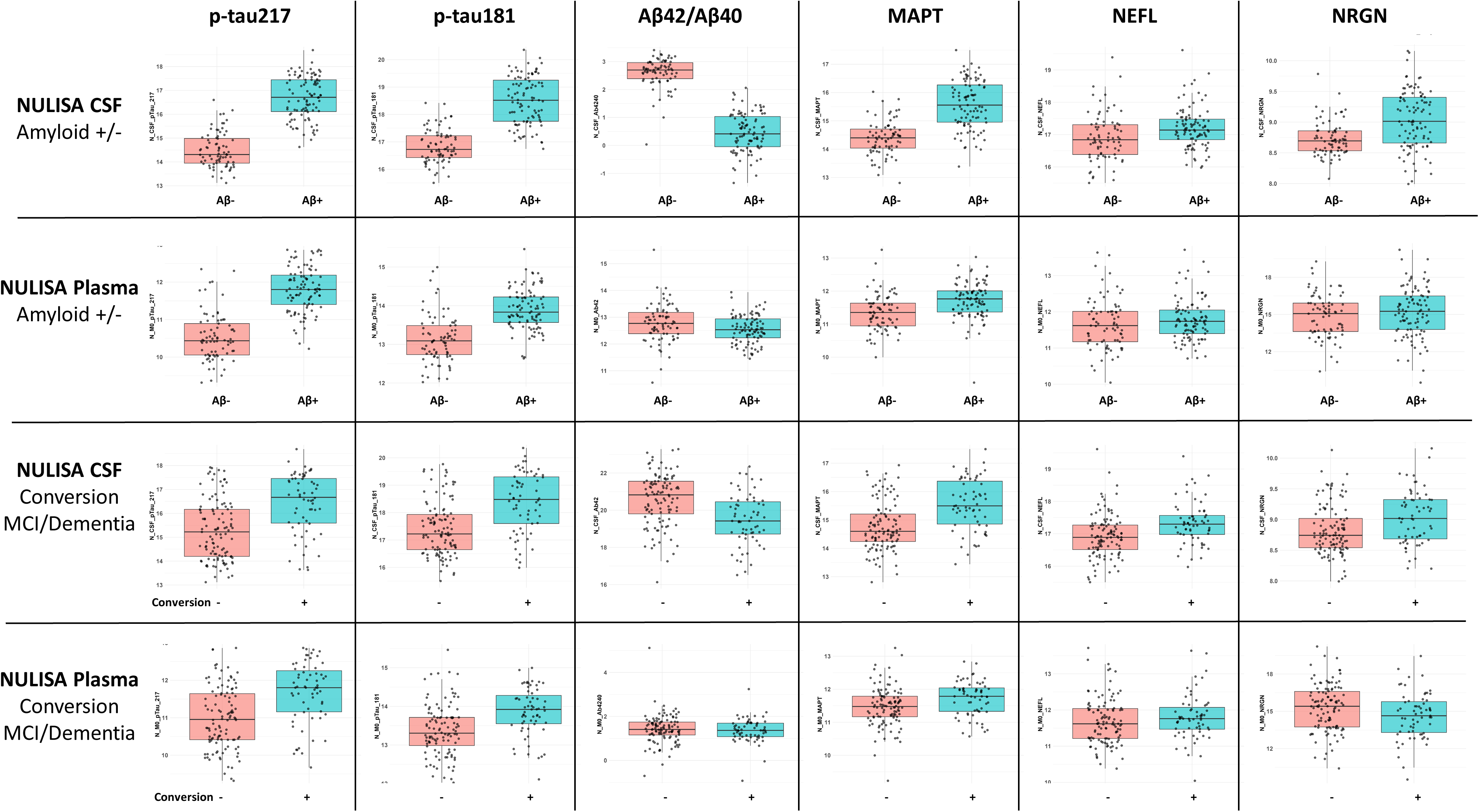
Distribution of CSF and plasma NULISA biomarkers according to amyloid +/-status and MCI-AD conversion +/-. Boxplots illustrating CSF and plasma biomarker levels in amyloid-positive versus amyloid-negative subjects (upper panels) and in MCI converters versus non-converters (lower panels). Differences in protein levels according to amyloid and conversion status were all significant, as assessed by adjusted linear mixed models controlling for age, sex, and APOE ε4, with p-values reported in Supplementary Table 3.

### Individual CSF and plasma biomarkers in MCI converters

As illustrated in the volcano plots of *Figures 3C and D* and detailed in *Supplementary Table 3*, the tau-related biomarkers most strongly associated with Aβ+ status were also the ones that differed most between converters and non-converters. However, CSF biomarkers linked to tau pathology (p-tau181) and to neurodegenerative processes ( MAPT, NEFL, or UCLH1) were also differential, with p-tau181 and MAPT matching p-tau217 performance. Amyloid biomarkers in the CSF also showed clear differential patterns. Considering the p-values without adjustment for multiple comparisons, we observed an increase in TREM1, which is involved in the microglial response. For synaptic-related biomarkers, we observed a mixed situation: SNCB, NRGN, and GDI1 were increased in CSF, but the latter two were decreased in plasma. We also observed that total and phosphorylated TDP-43 (TARDBP, pTDP43_409) were decreased in plasma converters. FGF2, which is important in neurogenesis, was decreased in plasma, as was RUVBL2, an actor in DNA repair. UBB and PDLIM5 also showed a small increase in CSF. Boxplots illustrating the distribution between converters and non-converters are shown in *Figure 4*, lower part.

### Combination of NULISA biomarkers for the detection of the conversion from MCI to dementia stage

The combination of biomarkers was performed using an elastic net logistic regression framework (see Methods). The ten most frequently observed combinations of two or three biomarkers were retained, and their corresponding AUCs were re-evaluated using 300 bootstrap replicates to estimate the AUC with 95% confidence intervals, along with model coefficients (*Supplementary Table 4*). Volcano plots of the best combination of CSF and plasma NULISA biomarkers, along with the ten best-performing individual AUCs, are shown in *Figures 5A and B*. Data including the logistic regression parameters are presented in *Table 2* and fully detailed in *Supplementary Table 4*. In both CSF and plasma, significant AUCs of individual biomarkers were below 0.75 (blue dots). In CSF, Aβ42, p-tau, as well as biomarkers linked to neuronal damage and neurodegeneration (MAPT, NEFL, UCLH1) had significant AUCs. In plasma, only p-tau biomarkers were significant.

**Figure 5:**
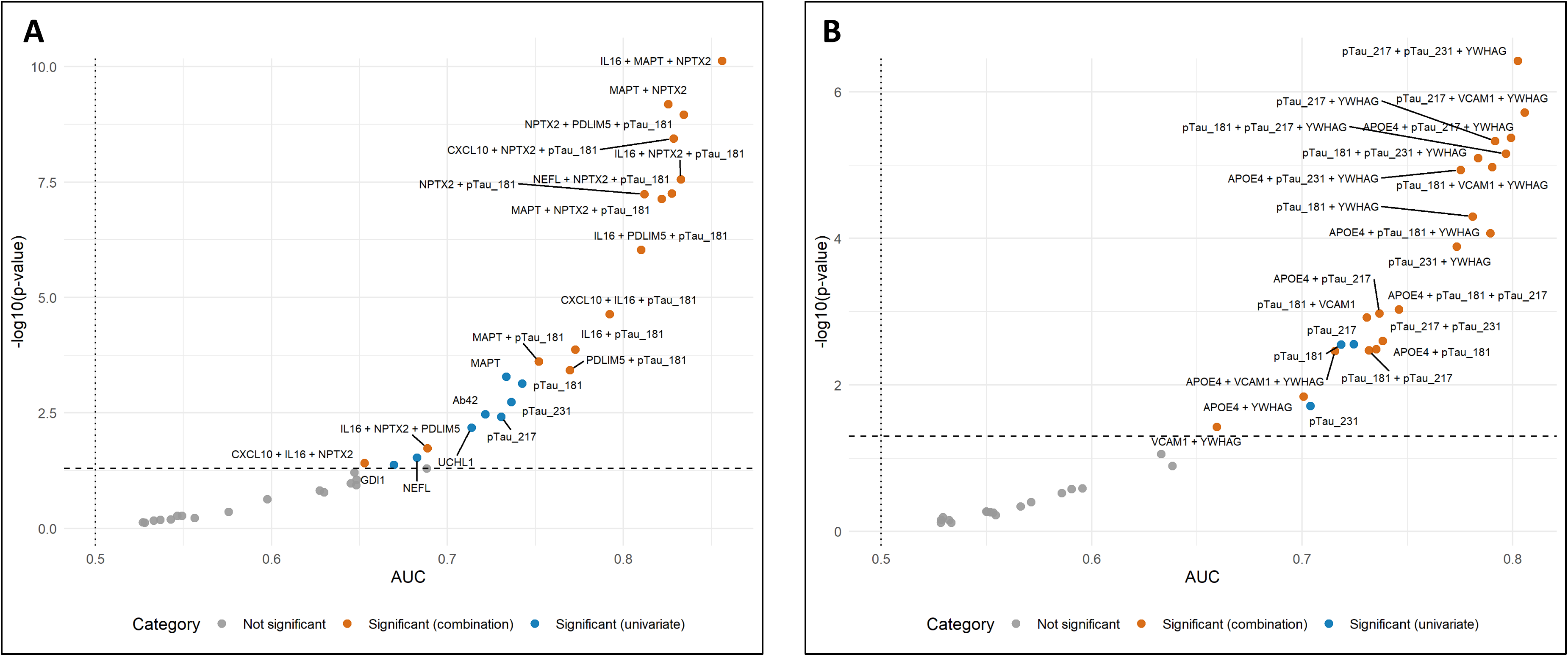
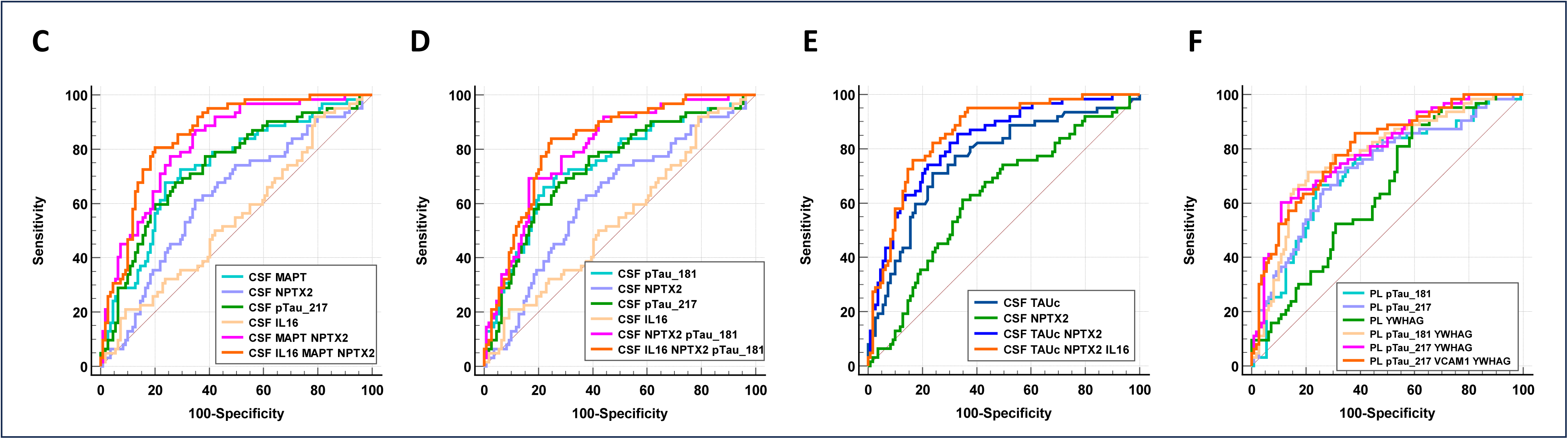
AUCs for conversion detection of single and combined NULISA biomarkers. Panels A and B: Volcano plots −log10(p-value) versus areas under the curve (AUCs) illustrating the performance of CSF (A) and plasma (B) NULISA biomarkers for MCI conversion detection. The limit for false discovery rate of 0.05 is indicated by the dotted line. Blue dots are associated with significant simplex biomarkers, and orange dots with regression combination of two or three. Panels C and D: receiver operating characteristic (ROC) curves for the following NULISA CSF biomarkers: MAPT, NPTX2, IL-16, pTau181, pTau217 and their combinations. Pairwise DeLong tests are significant for the following comparison: panel C : MAPT vs MAPT/NPTX2 (*p*=0.0042); MAPT vs IL-16 (*p*=0.0105); MAPT vs IL-16/MAPT/NPTX2 (*p*=0.0004); NPTX2 vs MAPT/NPTX2 (*p*=0.0001); NPTX2 vs IL-16/MAPT/NPTX2 (*p*<0.0001); MAPT/NPTX2 vs IL-16 (*p*<0.0001); IL-16 vs IL-16/MAPT/NPTX2 (*p*<0.0001), panel D : pTau181 vs IL-16 (*p*=0.0059); pTau181 vs pTau181/NPTX2 (*p*=0.0213); pTau181 vs pTau181/NPTX2/IL-16 (*p*=0.0050); NPTX2 vs pTau181/NPTX2 (*p*=0.0004); NPTX2 vs pTau181/NPTX2/IL-16 (*p*=0.0001); pTau217 vs IL-16 (*p*=0.0082); pTau217 vs pTau181/NPTX2 (*p*=0.0092); pTau217 vs pTau181/NPTX2/IL-16 (*p*=0.0024); IL-16 vs pTau181/NPTX2 (*p*=0.0001); IL-16 vs pTau181/NPTX2/IL-16 (*p*<0.0001). Panels E: ROC curves for the following CSF biomarkers and models: Lumipulse Tau (TAUc), NULISA NPTX2 and IL-16. Pairwise DeLong tests are significant for the following comparison: TAUc vs TAUc /NPTX2 (p=0.0249); TAUc vs NPTX2 (p=0.0036); TAUc vs TAUc /NPTX2 / IL16 (p= 0.0028); NPTX2 vs TAUc/NPTX2 (p<0.0001); NPTX2 vs TAUc/NPTX2/IL6 (p < 0.0001). Panel F: ROC curves for the following plasma NULISA biomarkers and models: p-tau181, p-tau217, YWHAG, VCAM1, and their combinations. Pairwise DeLong test are significant for the following comparison : p-tau217 vs VCAM1 (*p*=0.0058); pTau217 vs p-tau217/YWHAG (*p*=0.0158); VCAM1 vs p-tau181 (*p*=0.0072); VCAM1 vs p-tau217/YWHAG (*p*=0.0001); VCAM1 vs p-tau217/VCAM1 (*p*<0.0001); YWHAG vs p-tau217/YWHAG (*p*=0.0005); p-tau181 vs p-tau217/YWHAG (*p*=0.0169); p-tau217/YWHAG vs p-tau217/VCAM1 (*p*=0.0433). The numeric AUCs values along with the regression coefficients are reported in the table 2 and the supplementary table 4.

**Table 2:** Performance of CSF and plasma biomarkers and their combinations for prediction of MCI conversion.

| Fluid | Biomarkers | AUC | HR 1st vs 3rd | Regression coefficients |
| --- | --- | --- | --- | --- |
| CSF | IL16 / MAPT / NPTX2 | 0.86 (0.80-0.91) | 39.8 (9.6-165.2) | 8.9503 -1.6527*IL16 + 1.9436*MAPT -1.7068*NPTX2 |
| CSF | NPTX2 / PDLIM5 / p-tau181 | 0.83 (0.77-0.89) | 17.3 (6.1-48.8) | -16.3979 -1.4083*NPTX2 + 1.9152*PDLIM5 + 1.2287* p-tau181 |
| CSF | IL16 / NPTX2 / p-tau181 | 0.83 (0.77-0.89) | 15 (5.3-42.3) | 4.2311 -1.2308*IL16 -1.2185*NPTX2 + 1.3108* p-tau181 |
| CSF | MAPT / NPTX2 | 0.83 (0.76-0.88) | 15.8 (5.6-44.8) | -0.1981 + 1.5609*MAPT -1.6494*NPTX2 |
| CSF | NPTX2 / p-tau181 | 0.81 (0.75-0.87) | 13.7 (5.3-35.2) | -2.6105 -1.2526*NPTX2 + 1.1334* p-tau181 |
| CSF | IL16 / p-tau181 | 0.77 (0.70-0.84) | 6.4 (3.1-13.4) | -7.9988 -1.2926*IL16 + 1.0331* p-tau181 |
| CSF | p-tau181 | 0.74 (0.66-0.81) | 5.9 (2.9-12) | p-tau181 |
| CSF | MAPT | 0.73 (0.66-0.80) | 5.6 (2.8-11.4) | MAPT |
| CSF | p-tau217 | 0.73 (0.64-0.81) | 6.5 (3.1-13.6) | p-tau217 |
| CSF | Ab42 | 0.72 (0.63-0.80) | 4.8 (2.3-10.2) | Ab42 |
| CSF | NPTX2 | 0.63 (0.53-0.72) | 2 (1-3.7) | NPTX2 |
| CSF | IL16 | 0.54 (0.46-0.63) | 1.1 (0.6-2.1) | IL16 |
| Plasma | p-tau217 / VCAM1 / YWHAG | 0.81 (0.75-0.87) | 9.3 (4.1-21) | 40.7168 + 1.2379*p-tau217 -1.3334*VCAM1 -3.2667*YWHAG |
| Plasma | p-tau217 / YWHAG | 0.79 (0.73-0.86) | 7.4 (3.6-15.4) | 25.8546 + 1.1528*p-tau217 -3.454*YWHAG |
| Plasma | p-tau181 / YWHAG | 0.78 (0.70-0.85) | 9.4 (4.4-20.3) | 18.4307 + 1.3006*p-tau181 -3.2116*YWHAG |
| Plasma | p-tau217 | 0.72 (0.63-0.80) | 6.1 (2.9-12.8) | p-tau217 |
| Plasma | p-tau181 | 0.72 (0.64-0.79) | 5.4 (2.6-10.9) | p-tau181 |
| Plasma | YWHAG | 0.64 (0.47-0.73) | 2.6 (1.3-5.1) | YWHAG |
| Plasma | VCAM1 | 0.53 (0.45-0.64) | 1.5 (0.8-2.7) | VCAM1 |

**Legend:** The tables present the area under the curve (AUC, 95% confidence intervals) for each NULISA CSF and plasma biomarker or biomarker combinations, together with the corresponding logistic regression coefficients. Hazard ratios (HR) with 95% CI obtained between 1st vs. 3rd tercile of the biomarker-derived score are indicated.
Abbreviations: AUC, area under the receiver operating characteristic curve; CSF, cerebrospinal fluid; HRs, hazard ratios.

The CSF model with the highest performance combines IL16/MAPT/NPTX2 and reaches an AUC of 0.86 (0.80–0.91) (*Table 2*). The best model combining two biomarkers, MAPT/NPTX2, reaches an AUC of 0.83 (0.76–0.88). Visual illustration of the best AUCs generated with the CSF and plasma NULISA biomarkers, along with statistical comparison between the AUCs, are provided in *Figures 5 C,D and F*. Using the routine ELISA value of CSF tau (TAUc) instead of NULISA MAPT yielded the same improvement when combined with NPTX2 and IL16 to predict MCI conversion (*Figure 5E*). All the models including two or three CSF NULISA biomarkers with AUCs above 0.8 (*Table 2, Supplementary Table 4*) included both a neurodegenerative-related biomarker (MAPT) and the synaptic biomarker NPTX2. Importantly, the regression coefficient associated with NPTX2 is negative, indicating that its decrease provides valuable complementary value biomarkers to the increase observed for the neurodegenerative biomarker. The best-selected combinations of plasma biomarkers always combine p-tau biomarkers and YWHAG. Two p-tau biomarkers are sometimes present in the model, but the AUCs are only marginally improved (*Table 2, Supplementary Table 4*). APOE and VCAM1 are also well represented in the selected models. These results are illustrated by plotting ROC curves of selected biomarkers in *Figure 5F*.

### Survival curve and hazard risk for conversion from MCI to dementia stage

The best CSF and plasma biomarker models, based on their AUCs, were tested in a predictive survival Kaplan–Meier approach, where each biomarker was separated into terciles (see Methods). The log-rank test and hazard ratios (HRs) computed between the 1st and the 3rd tertiles are reported in *Table 2* and *Supplementary Table 4* and illustrated *<u>in</u> Figure 6*. CSF p-tau181, p-tau231, MAPT, p-tau217, and Aβ42 showed significant log-rank tests and HRs (95% CI) of 5.9 (2.9–12), 7.3 (3.4–15.9), 5.6 (2.8–11.4), 6.5 (3.1–13.6), and 4.8 (2.3–10.2), respectively. The combination of two CSF biomarkers showed an increase in HR with values of 15.8 (5.6–44.8), 13.7 (5.3–35.2), and 6.4 (3.1–13.4) for MAPT/NPTX2, NPTX2/p-tau181, or IL16/p-tau181. When three CSF biomarkers were combined, the best combination, IL16/MAPT/NPTX2, achieved a very high HR of 39.8 (9.6–165.2), while the other combinations, NPTX2/p-tau181/PDLIM5 and IL16/NPTX2/p-tau181, had lower HRs of 17.3 (6.1–48.8) and 15 (5.3–42.3), respectively.

**Figure 6.**
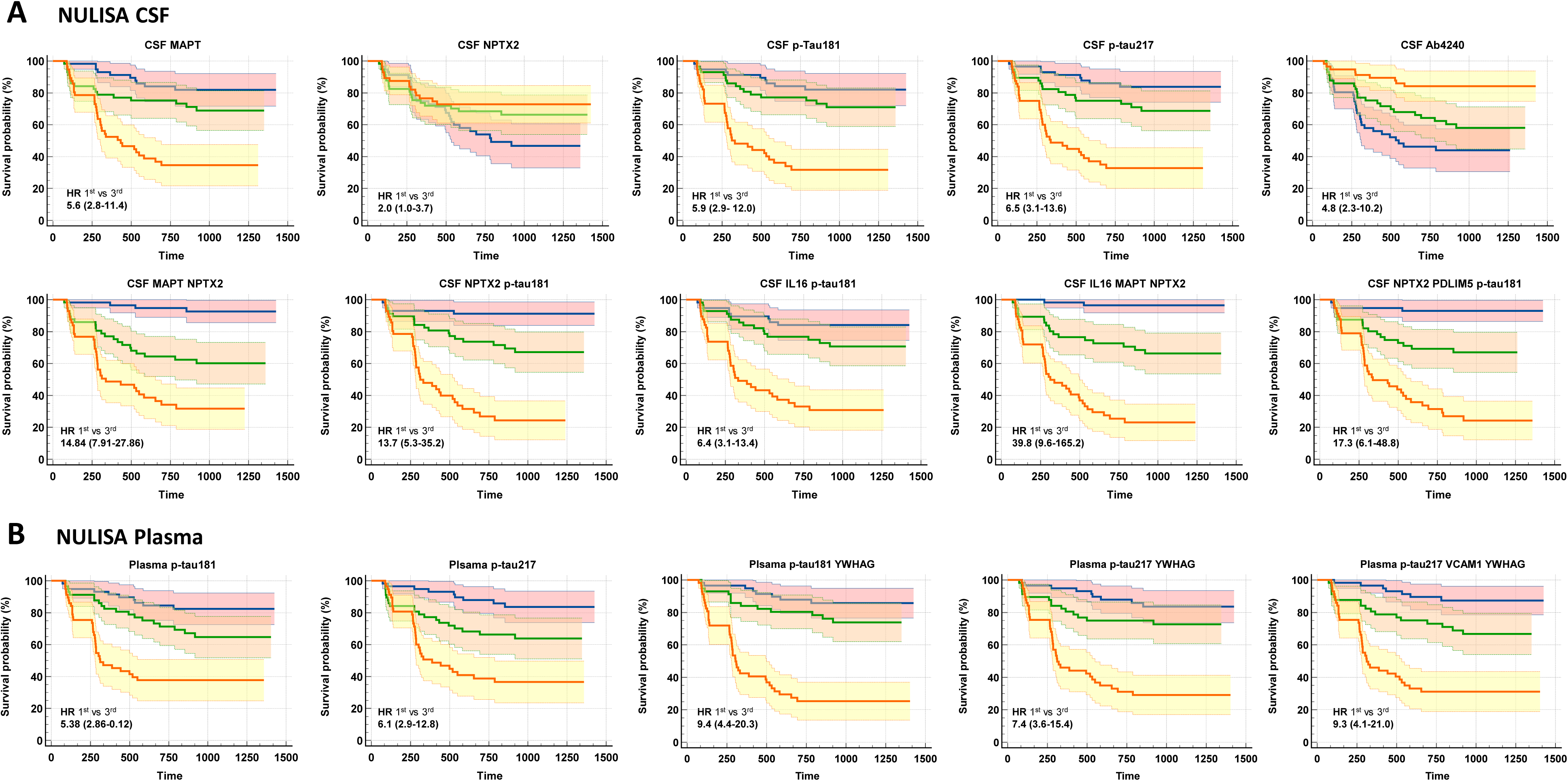
Survival analyses for NULISA biomarkers and their models. Kaplan–Meier curves for MCI conversion according to tertiles of selected NULISA CSF (A) and plasma (B) biomarkers and their combinations. Hazard ratios (HRs) comparing the 1st and 3rd tertiles are shown with 95% confidence intervals (CI). Regression coefficients, log-rank P values, and HRs with 95% CIs are reported in Table 2 and Supplementary Table 4.

In plasma, p-tau217 and p-tau181 alone reached HRs of 6.1 (2.9–12.8) and 5.4 (2.6–10.9), respectively. When combined with YWHAG (14-3-3γ), p-tau217 reached an HR of 7.4 (3.6–15.4) and p-tau181 an HR of 9.4 (4.4–20.3). Combining three plasma biomarkers did not improve this last HR.

## Discussion

In this work, we capitalized on the NULISA technology to evaluate the utility of a panel of CSF and plasma biomarkers. As with any new technology, ensuring both analytical reliability and biological relevance is essential. NULISA relies on NGS amplification of DNA oligonucleotides generated by immunocomplexes, and our results support its robustness based on the correlation with values obtained with other approaches on both CSF and plasma samples, in line with recent reports ^23, 24, 33^.

An important aspect of BALTAZAR is that plasma and CSF samples were collected on the same day, allowing a direct comparison of biomarker profiles across fluids. This parallel measurement provides insight into the capacity of each marker to be informative or not in blood. For example, p-tau species, which showed strong agreement between CSF and plasma are now recognized as robust blood biomarkers of AD ^10, 13, 34^. Conversely, other biomarkers, including synaptic proteins like NRGN or SNAP25, were not correlated between fluids, and it is noteworthy that they are informative in CSF but not in plasma ^35, 36^. These discrepancies between fluids likely arise from multiple factors, including 1) physiological differences in biomarker production between the CNS and peripheral compartments (as shown for ApoE), 2) the selectivity of blood–brain barrier transfer, and 3) potential analytical challenges related to protein abundance and assay sensitivity in blood.

In BALTAZAR, amyloid positivity (Aβ+) was defined using the CSF Aβ42/40 ratio measured with routine ELISA assays. Individual CSF and plasma NULISA targets associated with Aβ+ were represented primarily by amyloid and tau biomarkers. The relative performance of these biomarkers (p-tau217 > p-tau231 > p-tau181 > GFAP …) is fully concordant with previous observations in BALTAZAR ^13, 37^ and other cohorts head ^38, 39^, confirming the relevance and performance of NULISA detection. In CSF, additional associations were observed for markers of neuronal injury and synaptic function (e.g., NRGN, VSNL1, SNCA, UCHL1, NEFL), as well as astrocytic (GFAP) and immune-related proteins (CHIT1, SMOC1). This biomarker profile aligns closely with the current AD pathophysiology model, in which amyloid and tau initiate neuronal injury and synaptic dysfunction, while activated glial/microglial cells contribute to neuroinflammation and neurovascular alterations ^40^.

The detection of Aβ+ with plasma p-tau217 alone, or in combination with Aβ42, achieves already a very high level of correspondence with CSF or PET detection of Aβ+ that allows these biomarkers to be used at an individual level and in routine clinical practice ^41–43^. The BALTAZAR cohort offers a unique opportunity to study cognitive decline, blood biomarkers, and the conversion of MCI to dementia stage, a key clinical question. Notably, this cohort, composed of relatively older participants, showed a high conversion rate of 40% over three years of follow-up. Previous work within this cohort has explored the predictive value for amyloid positivity of different plasma biomarkers, including neurofilaments, amyloid peptides, p-tau181, and p-tau217 ^13, 21, 27, 32^. While p-tau217 performs well for identifying amyloid positivity, its ability to predict conversion to dementia stage is limited, with AUC values below 0.75 and sensitivity/specificity under 75%, therefore rendering its use for individual patient management difficult.

In our study, an important observation emerged first from the analysis of CSF biomarkers predictive of conversion. While CSF p-tau217, as expected, was among the strongest performers, markers of neurodegeneration such as tau (MAPT), NfL (NEFL), and UCHL1 performed equally well. CSF p-tau181, which may be considered both an N+ and a T+ biomarker within the AT/N framework ^9^, showed predictive value for conversion comparable to p-tau217. Synaptic biomarkers such as neurogranin (NRGN) and beta-synuclein (SNCB) were also increased in CSF, consistent with previous reports of their dynamic changes across the course of AD ^9, 44, 45^. In plasma, only p-tau217, p-tau181, and, at lower performance, YWHAG (14-3-3γ) remained significant after adjustment for conversion prediction. Interestingly, YWHAG was not differential in CSF, in contrast to previous studies using mass spectrometry ^46, 47^. This discrepancy may relate to methodological differences, as other recent NULISA studies also failed to identify YWHAG as associated with amyloid pathology ^23, 24^. YWHAG corresponds to the gamma isoform of the 14-3-3 protein, which was identified years ago as a surrogate marker of clinical utility in Creutzfeldt–Jakob disease ^48, 49^. Prion diseases, which represent neurodegeneration in the absence of amyloid pathology, highlight how biomarkers such as 14-3-3, tau, p-tau181, and synuclein can reflect neurodegenerative damages ^8, 50^.

The hazard ratios reported for the NULISA biomarkers were intended to compare biomarkers in order to identify the most informative ones. However, these HR values cannot be directly extrapolated to clinical use, as the analysis evaluated biomarkers as continuous variables by comparing the first and third tertiles in the cohort, thereby excluding the middle tertile from the calculation and without selecting a specific cutoff. In the current clinical routine, it is worth noting that amyloid status assessed using CSF already shows a significant hazard ratio of 3.2 (95% CI 1.9–5.0) for conversion to dementia and represents an asset for patient management.

Taken together, these results suggest that the mechanisms driving conversion from MCI to dementia stage are not exclusively linked to amyloid pathology but rather reflect broader processes of neurodegeneration and synaptic dysfunction. Synaptic biomarkers, in particular, have long attracted interest for their predictive value and their capacity to reflect disease dynamics over time ^51, 52^.

The interest of a multimarker approach lies in its ability to combine biomarkers to enhance prediction of conversion to dementia stage, while simultaneously providing insight into the underlying pathophysiological processes. In this study, biomarker combinations were restricted to a maximum of three, to minimize the risk of overfitting, following the “10 events per variable” rule for stable estimates ^53^. Increasing the number of biomarkers to four or five did not yield meaningful gains in detection accuracy and would impair translation to effective clinical chemistry methods.

Overall, combinations improved conversion prediction in both CSF and plasma, although performance remained higher in CSF. The best CSF model (IL16, MAPT, NPTX2) achieved an AUC of 0.86 and a striking hazard ratio of nearly 40 when comparing terciles. While this highlights the strong discriminatory capacity of this biomarker combination, such large effect sizes should be interpreted cautiously, as they may partly reflect model instability or overfitting in the context of a single cohort, despite the use of bootstrap validation to improve the robustness of the estimates. Even models such as MAPT with NPTX2 maintained strong predictive power (AUC 0.83), suggesting that two-biomarker panels may already provide substantial prognostic value.

These findings highlight the central role of neurodegenerative biomarkers (tau (MAPT), p-tau181, Nfl (NEFL)) as the “backbone” of conversion prediction, complemented by synaptic markers such as NPTX2. Notably, NPTX2 outperformed other synaptic candidates and has been repeatedly linked to AD progression in large-scale proteomics ^46, 47^. The selection of IL6 and CXCL10 in our CSF models is also of interest, as both cytokines are implicated in microglial and T-lymphocyte recruitment to the CNS ^54, 55^. Microglia may initially play a protective role by clearing amyloid ^56^, but chronic activation can promote neuronal injury and thereby contribute to the conversion process. Similarly, T-cell infiltration, facilitated by blood–brain barrier dysfunction, is increasingly recognized as a contributor to neuroinflammation in AD ^57^.

In plasma, the best combinations for conversion prediction consistently associate p-tau181 or p-tau217 with YWHAG (14-3-3γ), reminding the CSF situation with markers of neurodegeneration. VCAM1 also appeared in some models, consistent with its reported role in microglial amyloid clearance ^58^, although its added predictive value was limited. Importantly, unlike in CSF, no synaptic biomarker improved the plasma models for conversion prediction. This finding is consistent with our observation that synaptic markers did not correlate between CSF and plasma.

Conversion to dementia was markedly higher in the Aβ+ group than in the Aβ− group, although the latter still reached a rate of 20%. Examination of the biological profiles of these participants showed that approximately half were borderline, with either pathological CSF p-tau181 levels and/or an Aβ42/40 ratio close (within <10%) to the cutoff. Unfortunately, a second CSF sample was not available. Overall, this suggests that a small proportion of conversions (<10%) occurred without biological evidence supporting AD. These participants may partly account for the contribution of non-specific AD biomarker profiles to the observed conversions. One limitation of our study is that it is observational and restricted to the measurement of biomarker concentrations in CSF and plasma, without anatomopathological confirmation. In addition, there is an inherent risk of circular reasoning when exploring determinants of Aβ+ in the BALTAZAR cohort, since CSF biomarkers were both used for classification and analyzed for variation. Importantly, however, the central focus and main conclusions of this work relate to conversion to dementia stage, which was determined independently of biomarker status. Another limitation is the reliance on a single cohort, which could introduce specific biases. Nonetheless, the BALTAZAR cohort has previously been used to investigate the value and limitations of blood biomarkers for AD ^9, 13, 21, 27, 32, 37, 59–61^, and conclusions from these studies have been reproduced in other cohorts, supporting the reliability of this dataset.

In conclusion, combinations of CSF biomarkers can predict conversion of MCI patients to dementia stage with high accuracy. The strongest associations involved total tau (measured either with NULISA or routine assays) and NPTX2, with additional improvement when neuroinflammatory markers are included. In plasma, however, the translation of this signature seems less optimal: the best-performing combination, p-tau181 with gamma 14-3-3 (YWHAG), remained markedly less predictive.

Beyond their predictive value, these findings also carry important pathophysiological implications. They suggest that the processes driving conversion from MCI to dementia stage are not determined solely by the core amyloid and tau biomarkers, but rather by downstream neurodegenerative and synaptic alterations, complemented by inflammatory responses. This reinforces the idea that biomarkers of neuronal and synaptic injury may be crucial for capturing disease progression and for improving patient stratification in clinical practice and therapeutical trials.

## Declarations

### Ethics approval and consent to participate

Written informed consent to participate in the study was provided by all participants. The BALTAZAR study has approval from by the Paris ethics committee under # 2010-A00335-34 (CPP Ile de France IV Saint-Louis Hospital). The protocol is registered on ClinicalTrial under number NCT01315639.

### Availability of data and materials

Data and informed consent form are available upon request after publication (APHP, Paris). Requests will be considered by each study investigator, based on the information provided by the requester, regarding the study and analysis plan. If the use is appropriate, a data sharing agreement will be put in place before distributing a fully de-identified version of the dataset, including the data dictionary used for analysis with individual participant data.

### BALTAZAR study group

The BALTAZAR study group: Olivier Hanon [1], Frédéric Blanc [2], Yasmina Boudali [1], Audrey Gabelle [3], Jacques Touchon [3], Marie- Laure Seux [1], Hermine Lenoir [1], Catherine Bayle [1], StéphanieBombois [4], Christine Delmaire [4], Xavier Delbeuck [5], Florence Moulin [1], Emmanuelle Duron [6], Florence Latour [7], Matthieu Plichart [1], Sophie Pichierri [8], Galdric Orvoën [1], Evelyne Galbrun [9], Giovanni Castelnovo [10], Lisette Volpe-Gillot [11], Florien Labourée [1], Pascaline Cassagnaud [12], Claire Paquet [13], Françoise Lala [14], Bruno Vellas [14], Julien Dumurgier [13], Anne-Sophie Rigaud [1], Christine Perret-Guillaume [15], Eliana Alonso [16], Foucaud du Boisgueheneuc [17], Laurence Hugonot-Diener [1], Adeline Rollin-Sillaire [12], Olivier Martinaud [18], Clémence Boully [1], Yann Spivac [19], Agnès Devendeville [20], Joël Belmin [21], Philippe Robert [22], Thierry Dantoine [23], Laure Caillard [1], David Wallon [24], Didier Hannequin [18], Nathalie Sastre [14], Sophie Haffen [25], Anna Kearney-Schwartz [15], Jean-Luc Novella [26], Vincent Deramecourt [12], Valérie Chauvire [27], Gabiel Abitbol [1], Nathalie Schwald [19], Caroline Hommet [28], François Sellal [29], Marie-Ange Cariot [16], Mohamed Abdellaoui [30], Sarah Benisty [31], Salim Gherabli [1], Pierre Anthony [29], Frédéric Bloch [32], Nathalie Charasz [1], Sophie Chauvelier [1], Jean-Yves Gaubert [1], Guillaume Sacco [22], Olivier Guerin [22], Jacques Boddaert [33], Marc Paccalin [17], Marie-Anne Mackowiak [12], Marie-Thérèse Rabus [9], Valérie Gissot [34], Athanase Benetos [15], Candice Picard [20], Céline Guillemaud [35], Gilles Berrut [8], Claire Gervais [22], Jacques Hugon [13], Jean-Marc Michel [29], Jean- Philippe David [19], Marion Paulin [12], Pierre-JeanOusset [14], Pierre Vandel [36], Sylvie Pariel [21], Vincent Camus [37], Anne Chawakilian [1], Léna Kermanac’h [1], Anne-Cécile Troussiere [12], Cécile Adam [23], Diane Dupuy [20], Elena Paillaud [16], Hélène Briault [9], Isabelle Saulnier [38], Karl Mondon [37], Marie-Agnès Picat [23], Marie Laurent [16], Olivier Godefroy [20], RezkiDaheb [16], Stéphanie Libercier [29], Djamila Krabchi [1], Marie Chupin [39], JeanSébastien Vidal [1], Edouard Chaussade [1], Christiane Baret-Rose [40], Sylvain Lehmann [41], Bernadette Allinquant [40], Susanna Schraen-Maschke [4].

[1] EA 4468, APHP, Hospital Broca, Memory Resource and Research Centre of de Paris-Broca-Ile de France, Université Paris Cité, F-75013 Paris, France [2] CHRU de Strasbourg, Memory Resource and Research Centre of Strasbourg/Colmar, French National Centre for Scientific Research, ICube Laboratory and Fédération de Médecine Translationnelle de Strasbourg, Team Imagerie Multimodale Intégrative en Santé /Neurocrypto, Université de Strasbourg, F-67000 Strasbourg, France [3] Memory Research and Resources Center, Department of Neurology, Inserm INM NeuroPEPs Team, Université de Montpellier, F-34000 Montpellier, France [4] Inserm, CHU Lille, UMR-S-U1172, LiCEND, Lille Neuroscience & Cognition, LabEx DISTALZ, University of Lille, F-59000 Lille, France [5] Univ. Lille, Inserm U1171 Degenerative and Vascular Cognitive Disorders, F-59000 Lille, France. [6] Université Paris-Saclay, APHP, Hôpital Paul Brousse, département de gériatrie, Équipe MOODS, Inserm 1178, F-94800 Villejuif, France. [7] Centre Hospitalier de la Côte Basque, Department of Gerontology, F-64100 Bayonne, France. [8] Université de Nantes, EA 4334 Movement-Interactions-Performance, CHU Nantes, Memory Research Resource Center of Nantes, Department of clinical gerontology, F-44000 Nantes, France. [9] Sorbonne Université, APHP, Centre Hospitalier Dupuytren, Department of Gérontology 2, F-91210 Draveil, France. [10] CHU de Nimes, Hôpital Caremeau, Neurology Department, F- 30029 Nimes, France. [11] Hôpital Léopold Bellan, Service de Neuro-Psycho-Gériatrie, Memory Clinic, F-75014 Paris, France. [12] Univ. Lille, CHU de Lille, Memory Resource and Research Centre of Lille, Department of Neurology, F-59000 Lille, France. [13] GHU APHP Nord Lariboisière Fernand Widal, Centre de Neurologie Cognitive, Université Paris Cité, F-75010 Paris, France [14] Université de Toulouse III, CHU La Grave-Casselardit, Memory Resource and Research Centre of Midi-Pyrénées, F-31300 Toulouse, France. [15] Université de Lorraine, CHRUdeNancy, Memory Resource and Research Centre of Lorraine, F-54500 Vandoeuvre-lès-Nancy, France. [16] Université de Paris, APHP, Hôpital européen Georges Pompidou, Service de Gériatrie, F-75015, Paris, France. [17] CHU de Poitiers, Memory Resource and Research Centre of Poitiers, F-86000 Poitiers, France. [18] CHU Charles Nicolle, Memory Resource and Research Centre of Haute Normandie, F-76000 Rouen, France. [19] APHP, Centre Hospitalier Émile-Roux, Department of Gérontology 1, F-94450 Limeil-Brévannes, France. [20] CHU d’Amiens-Picardie, Memory Resource and Research Centre of Amiens Picardie, F-80000 Amiens, France. [21] Sorbonne Université, APHP, Hôpitaux Universitaires Pitie- Salpêtrière-Charles-Foix, Service de Gériatrie Ambulatoire, F-75013 Paris, France. [22] Université Côte d’Azur, CHU de Nice, Memory Research Resource Center of Nice, CoBTek lab, F-06100 Nice, France. [23] CHU de Limoges, Memory Research Resource Center of Limoges, F-87000 Limoges, France. [24] Normandie Univ, UNIROUEN, Inserm U1245, CHU de Rouen, Department of Neurology and CNR-MAJ, Normandy Center for Genomic and Personalized Medicine, CIC-CRB1404, F-76000, Rouen, France. [25] CHU de Besançon, Memory Resource and Research Centre of Besançon Franche-Comté, F-25000 Besançon, France. [26] Université de Reims Champagne-Ardenne, EA 3797, CHU de Reims, Memory Resource and Research Centre of Champagne- Ardenne, F-51100 Reims, France. [27] CHU d’Angers, Memory Resource and Research Centre of Angers, F-49000 Angers, France. [28] CHRU de Tours, Memory Resource and Research Centre of Tours, F-37000 Tours, France. [29] Université de Strasbourg, CHRU de Strasbourg, Memory Resource and Research Centre of Strasbourg/Colmar, Inserm U-118, F-67000 Strasbourg, France. [30] Univ Paris Est Creteil, EA 4391 Excitabilité Nerveuse et Thérapeutique, CHU Henri Mondor, Department of Neurology, F- 94000 Créteil, France. [31] Hôpital Fondation Rothschild, Department of Neurology, F- 75019 Paris, France. [32] CHU d’Amiens-Picardie, Department of Gerontology, F-80000 Amiens, France. [33] Sorbonne Université, APHP, Hôpitaux Universitaires Pitie- Salpêtrière-Charles Foix, Memory Resource and Research Centre, Centre des Maladies Cognitives et Comportementales IM2A, Inserm UMR 8256, F-75013 Paris, France. [34] Université François-Rabelais de Tours, CHRU de Tours, MemoryResource andResearchCentre of Tours, Inserm CIC 1415, F-37000 Tours, France. [35] Sorbonne Université, APHP, Hôpitaux Universitaires Pitie- Salpêtrière-Charles Foix, Memory Resource and Research Centre, Centre des Maladies Cognitives et Comportementales IM2A, F-75013 Paris, France. [36] Université Bourgogne Franche-Comté, Laboratoire de Recherches Intégratives en Neurosciences et Psychologie Cognitive, CHU de Besançon, Memory Resource and Research Centre of Besançon Franche-Comté, F-25000 Besançon, France. [37] Université François-Rabelais de Tours, CHRU de Tours, UMR Inserm U1253, F-37000 Tours, France. [38] Université de Limoges, EA 6310 HAVAE, CHU de Limoges, Memory Research Resource Center of Limoges, F-87000 Limoges, France. [39] Université Paris-Saclay, Neurospin, CEA, CNRS, catineuroimaging.com, CATI Multicenter Neuroimaging Platform, F-91190 Gif-sur-Yvette, France. [40] Université de Paris, Institute of Psychiatric and Neurosciences, Inserm UMR-S 1266, F-75014 Paris, France. [41] Univ Montpellier, IRMB CHU de Montpellier, INM INSERM, Montpellier, France

## Authors’ contributions

S.L., J.S.V., CD and O.H. take responsibility for the integrity of the data and the accuracy of the data analysis.

Concept and design: O.H., S.B., A.G., S S-M., S.L., C.D.

Acquisition, analysis, or interpretation of data: All authors.

Drafting of the manuscript: S.L.

Critical revision of the manuscript for important intellectual content: All authors.

Statistical analysis: S.L., J.S.V.

Obtained funding: O.H., S.B., A.G., S S-M., C.P., C.H., S.L., C.D.

All authors had full access to the data and contributed to revision and editing of the manuscript.

