## Supplementary material for "Beyond amyloid and tau: synaptic and neurodegenerative biomarkers shape MCI progression"

**Supplementary Table 1:** List of biomarkers measured with the NULISA platform in CSF and plasma.

| **Uniprot ID** | **Gene** | **Full Gene Name** | **Protein Name** | **Common Name** | **NULISA** | **CSF detection** | **Plasma detection** |
| --- | --- | --- | --- | --- | --- | --- | --- |
| P05067 | Abeta38 | Amyloid Beta Precursor Protein | Amyloid-beta precursor protein | ab38 | Ab38 | 100% | 100% |
| P05067 | Abeta40 | Amyloid Beta Precursor Protein | Amyloid-beta precursor protein | ab40 | Ab40 | 100% | 100% |
| P05067 | Abeta42 | Amyloid Beta Precursor Protein | Amyloid-beta precursor protein | ab42 | Ab42 | 100% | 100% |
| P22303 | ACHE | Acetylocholinesterase (Cartwright Blood Group) | Acetylcholinesterase | ACHE | ACHE | 100% | 100% |
| O00468 | AGRN | Agrin | Agrin | AGRN | AGRN | 100% | 100% |
| P08758 | ANXA5 | Annexin A5 | Annexin A5 | ANXA5 | ANXA5 | 96% | 100% |
| P02649 | APOE | Apolipoprotein E | Apolipoprotein E | APOE | APOE | 100% | 100% |
| P02649 | APOE | Apolipoprotein E isoform 4 | Apolipoprotein E isoform 4 | APOE4 | APOE4* | 100% (E4) | 100% (E4) |
| P15289 | ARSA | Arylsulfatase A | Arylsulfatase A | ARSA | ARSA | 45% | 100% |
| P56817 | BACE1 | Beta-Secretase 1 | Beta-secretase 1 | BACE1 | BACE1 | 100% | 100% |
| P80723 | BASP1 | Brain Abundant Membrane Attached Signal Protein 1 | Brain acid soluble protein 1 | BASP1 | BASP1 | 100% | 100% |
| P23560 | BDNF | Brain Derived Neurotrophic Factor | Brain-derived neurotrophic factor | BDNF | BDNF | **8%** | 100% |
| P22676 | CALB2 | Calbindin 2 | Calretinin | CALB2 | CALB2 | 100% | 100% |
| P51671 | CCL11 | C-C Motif Chemokine Ligand 11 | Eotaxin | Eotaxin | CCL11 | 100% | 100% |
| Q99616 | CCL13 | C-C Motif Chemokine Ligand 13 | C-C motif chemokine 13 | MCP4 | CCL13 | 54% | 100% |
| Q92583 | CCL17 | C-C Motif Chemokine Ligand 17 | C-C motif chemokine 17 | TARC/CCL17 | CCL17 | **21%** | 100% |
| P13500 | CCL2 | C-C Motif Chemokine Ligand 2 | C-C motif chemokine 2 | MCP1 | CCL2 | 100% | 100% |
| O00626 | CCL22 | C-C Motif Chemokine Ligand 22 | C-C motif chemokine 22 | MDC | CCL22 | 41% | 100% |
| Q9Y258 | CCL26 | C-C Motif Chemokine Ligand 26 | C-C motif chemokine 26 | Eotaxin-3 | CCL26 | 100% | 100% |
| P10147 | CCL3 | C-C Motif Chemokine Ligand 3 | C-C motif chemokine 3 | MIP1a/CCL3 | CCL3 | 100% | 100% |
| P13236 | CCL4 | C-C Motif Chemokine Ligand 4 | C-C motif chemokine 4 | Mip1b/CCL4 | CCL4 | 98% | 100% |
| P29965 | CD40LG | CD40 Ligand | CD40 ligand | CD40L/TNFSF5 | CD40LG | **22%** | 100% |
| P08962 | CD63 | CD63 Molecule | CD63 antigen | CD63 | CD63 | 100% | 100% |
| P36222 | CHI3L1 | Chitinase 3 Like 1 | Chitinase-3-like protein 1 | YKL40 | CHI3L1 | 100% | 100% |
| Q13231 | CHIT1 | Chitinase 1 | Chitotriosidase-1 | CHIT1 | CHIT1 | 98% | 98% |
| Q02246 | CNTN2 | Contactin 2 | Contactin-2 | CNTN2 | CNTN2 | 100% | 87% |
| P06850 | CRH | Corticotropin Releasing Hormone | Corticoliberin | CRH | CRH | 100% | 100% |
| P02741 | CRP | C-Reactive Protein | C-reactive protein | CRP | CRP | 100% | 100% |
| P04141 | CSF2 | Colony Stimulating Factor 2 | Granulocyte-macrophage colony-stimulating factor | GM-CSF | CSF2 | 100% | 100% |
| P01034 | CST3 | Cystatin 3 | Cystatin-C | CST3 | CST3 | 100% | 100% |
| P78423 | CX3CL1 | C-X3-C Motif Chemokine Ligand 1 | Fractalkine | CX3CL1/Fractalkine | CX3CL1 | 100% | 100% |
| P09341 | CXCL1 | C-X-C Motif Chemokine Ligand 1 | Growth-regulated alpha protein | CXCL1/GROa | CXCL1 | 100% | 100% |
| P02778 | CXCL10 | C-X-C Motif Chemokine Ligand 10 | C-X-C motif chemokine 10 | IP-10 | CXCL10 | 100% | 100% |
| P10145 | CXCL8 | C-X-C Motif Chemokine Ligand 8 | Interleukin-8 | IL8 | CXCL8 | 100% | 100% |
| P20711 | DDC | Aromatic-L-Amino-Acid Decarboxylase | Aromatic-L-Amino-Acid Decarboxylase | DDC | DDC | 80% | 100% |
| P09104 | ENO2 | Enolase 2 | Gamma-enolase | ENO2 | ENO2 | 100% | 100% |
| P05413 | FABP3 | Fatty Acid Binding Protein 3 | Fatty acid-binding protein, heart | FABP3 | FABP3 | 100% | 100% |
| Q15485 | FCN2 | Ficolin 2 | Ficolin-2 | FCN2 | FCN2 | **3%** | 100% |
| P09038 | FGF2 | Fibroblast Growth Factor 2 | Fibroblast growth factor 2 | FGF basic | FGF2 | 100% | 100% |
| P17948 | FLT1 | Fms Related Receptor Tyrosine Kinase 1 | Vascular endothelial growth factor receptor 1 | VEGF R1 | FLT1 | 100% | 100% |
| P15328 | FOLR1 | Folate Receptor Alpha | Folate receptor alpha | FOLR1 | FOLR1 | 100% | 100% |
| Q99988 | GDF15 | Growth Differentiation Factor 15 | Growth/differentiation factor 15 | GDF15 | GDF15 | 94% | 94% |
| P31150 | GDI1 | Rab GDP Dissociation Inhibitor Alpha | Rab GDP dissociation inhibitor alpha | GDI1 | GDI1 | 100% | 100% |
| P39905 | GDNF | Glial Cell Derived Neurotrophic Factor | Glial cell line-derived neurotrophic factor | GDNF | GDNF | 100% | 100% |
| P14136 | GFAP | Glial Fibrillary Acidic Protein | Glial fibrillary acidic protein | GFAP | GFAP | 100% | 100% |
| P17174 | GOT1 | Glutamic-Oxaloacetic Transaminase 1 | Aspartate aminotransferase, cytoplasmic | GOT1 | GOT1 | 100% | 100% |
| P69905 | HBA1; HBA2 | Hemoglobin Subunit Alpha 1 | Hemoglobin subunit alpha 1 \|Hemoglobin subunit alpha 2 | HBα1; HBα2 | HBA1 | **22%** | 76% |
| P42858 | HTT | Huntingtin | Huntingtin | HTT | HTT | 100% | 100% |
| P05362 | ICAM1 | Intercellular Adhesion Molecule 1 | Intercellular adhesion molecule 1 | ICAM1 | ICAM1 | 44% | 100% |
| P01579 | IFNG | Interferon Gamma | Interferon gamma | IFN-gamma | IFNG | 100% | 100% |
| P08069 | IGF1R | Insulin Like Growth Factor 1 Receptor | Insulin-like growth factor 1 Receptor | IGF1R | IGF1R | 100% | 100% |
| Q16270 | IGFBP7 | Insulin Like Growth Factor Binding Protein 7 | Insulin-like growth factor-binding protein 7 | IGFBP7 | IGFBP7 | 100% | 100% |
| P22301 | IL10 | Interleukin 10 | Interleukin-10 | IL-10 | IL10 | 100% | 100% |
| P29459\|P29460 | IL12A\|IL12B | Interleukin 12A \| Interleukin 12B | Interleukin 12A \| Interleukin 12B | IL-12p70 | IL12p70 | 100% | 100% |
| P35225 | IL13 | Interleukin 13 | Interleukin-13 | IL-13 | IL13 | 100% | 100% |
| P40933 | IL15 | Interleukin 15 | Interleukin-15 | IL-15 | IL15 | 100% | 100% |
| Q14005 | IL16 | Interleukin 16 | Pro-interleukin-16 | IL-16 | IL16 | 100% | 100% |
| Q16552 | IL17A | Interleukin 17A | Interleukin-17A | IL-17A | IL17A | 100% | 100% |
| Q14116 | IL18 | Interleukin 18 | Interleukin-18 | IL-18 | IL18 | 85% | 100% |
| P01584 | IL1B | Interleukin 1 Beta | Interleukin-1 beta | IL-1 beta | IL1B | 100% | 100% |
| P60568 | IL2 | Interleukin 2 | Interleukin-2 | IL-2 | IL2 | 91% | 100% |
| O95760 | IL33 | Interleukin 33 | Interleukin-33 | IL-33 | IL33 | 100% | 100% |
| P05112 | IL4 | Interleukin 4 | Interleukin-4 | IL-4 | IL4 | 100% | 100% |
| P05113 | IL5 | Interleukin 5 | Interleukin-5 | IL-5 | IL5 | 100% | 100% |
| P05231 | IL6 | Interleukin 6 | Interleukin-6 | IL-6 | IL6 | 100% | 100% |
| P08887 | IL6R | Interleukin 6 Receptor | Interleukin-6 receptor subunit alpha | IL-6R a | IL6R | 89% | 100% |
| P13232 | IL7 | Interleukin 7 | Interleukin-7 | IL-7 | IL7 | 100% | 100% |
| P15248 | IL9 | Interleukin 9 | Interleukin-9 | IL-9 | IL9 | 100% | 100% |
| P35968 | KDR | Kinase Insert Domain Receptor | Vascular endothelial growth factor receptor 2 | VEGF R2 | KDR | 92% | 98% |
| Q92876 | KLK6 | Kallikrein Related Peptidase 6 | Kallikrein-6 | KLK6 | KLK6 | 100% | 100% |
| P10636 | MAPT | Microtuble Associated Protein Tau | Brain-derived Tau | BD-MAPT | MAPT | 100% | 100% |
| P40925 | MDH1 | Malate Dehydrogenase 1 | Malate dehydrogenase, cytoplasmic | MDH1 | MDH1 | 100% | 100% |
| P08473 | MME | Membrane Metalloendopeptidase | Neprilysin | MME | MME | **1%** | 100% |
| Q13421 | MSLN | Mesothelin | Mesothelin | MSLN | MSLN | 100% | 100% |
| P12036 | NEFH | Neurofilament Heavy Chain | Neurofilament heavy polypeptide | NEFH | NEFH | 100% | 100% |
| P07196 | NEFL | Neurofilament Light Chain | Neurofilament light polypeptide | NfL | NEFL | 100% | 100% |
| P01138 | NGF | Nerve Growth Factor | Beta-nerve growth factor | NGF | NGF | 52% | 100% |
| Q15818 | NPTX1 | Neuronal Pentraxin 1 | Neuronal pentraxin-1 | NPTX1 | NPTX1 | 100% | 100% |
| P47972 | NPTX2 | Neuronal Pentraxin 2 | Neuronal pentraxin-2 | NPTX2 | NPTX2 | 100% | 100% |
| O95502 | NPTXR | Neuronal Pentraxin Receptor | Neuronal pentraxin receptor | NPTXR | NPTXR | 100% | 100% |
| P01303 | NPY | Neuropeptide Y | Pro-neuropeptide Y | NPY | NPY | 100% | 100% |
| Q92686 | NRGN | Neurogranin | Neurogranin | NRGN | NRGN | 100% | 100% |
| P37840 | Oligo-SNCA | Synuclein Alpha | Alpha-synuclein | SNCAagg | Oligo-SNCA | 100% | 100% |
| Q99497 | PARK7 | Parkinsonism Associated Deglycase | Parkinson disease protein 7 | PARK7 | PARK7 | 100% | 100% |
| P09619 | PDGFRB | Platelet Derived Growth Factor Receptor Beta | Platelet-derived growth factor receptor beta | PDGFRB | PDGFRB | 98% | 100% |
| Q96HC4 | PDLIM5 | PDZ And LIM Domain 5 | PDZ and LIM domain protein 5 | PDLIM5 | PDLIM5 | 100% | 100% |
| P49763 | PGF | Placental Growth Factor | Placenta growth factor | PLGF | PGF | 100% | 100% |
| P00558 | PGK1 | Phosphoglycerate Kinase 1 | Phosphoglycerate kinase 1 | PGK1 | PGK1 | **2%** | 100% |
| Q15063 | POSTN | Periostin | Periostin | POSTN | POSTN | **33%** | 100% |
| P30041 | PRDX6 | Peroxiredoxin 6 | Peroxiredoxin-6 | PRDX6 | PRDX6 | 100% | 100% |
| P49768 | PSEN1 | Presenilin 1 | Presenilin-1 | PSEN1 | PSEN1 | 100% | 100% |
| P37840 | SNCA | Phosphorylated alpha-synuclein | Phosphorylated alpha-synuclein at serine 129 | pSNCA-129 | pSNCA-129 | 100% | 100% |
| P10636 | pTau181 | Microtubule Associated Protein Tau | Microtubule-associated protein tau | pTau181 | pTau-181 | 100% | 100% |
| P10636 | pTau217 | Microtubule Associated Protein Tau | Microtubule-associated protein tau | pTau217 | pTau-217 | 100% | 100% |
| P10636 | pTau231 | Microtubule Associated Protein Tau | Microtubule-associated protein tau | pTau231 | pTau-231 | 100% | 100% |
| Q13148 | TARDBP | TAR DNA Binding Protein | TDP-43 with phosphorylation on serine 409 | pTDP43-409 | pTDP43-409 | 100% | 100% |
| P21246 | PTN | Pleiotrophin | Pleiotrophin | PTN | PTN | 100% | **3%** |
| Q13127 | REST | RE1 Silencing Transcription Factor | RE1-silencing transcription factor | REST | REST | 100% | 100% |
| Q9Y230 | RUVBL2 | RuvB Like AAA ATPase 2 | RuvB-like 2 | RUVBL2 | RUVBL2 | 100% | 100% |
| P80511 | S100A12 | S100 Calcium Binding Protein A12 | Protein S100-A12 | S100A12 | S100A12 | 16% | 100% |
| P04271 | S100B | S100 Calcium Binding Protein B | Protein S100-B | S100B | S100B | 100% | 100% |
| P0DJI8 | SAA1 | Serum Amyloid A1 | Serum amyloid A-1 protein | SAA1 | SAA1 | 3% | 100% |
| Q8N474 | SFRP1 | Secreted Frizzled Related Protein 1 | Secreted frizzled-related protein 1 | SFRP1 | SFRP1 | 100% | 100% |
| P35247 | SFTPD | Pulmonary Surfactant-Associated Protein | Pulmonary surfactant-associated protein D | SFTPD | SFTPD | 99% | 100% |
| O94813 | SLIT2 | Slit Guidance Ligand 2 | Slit homolog 2 protein | SLIT2 | SLIT2 | 100% | 100% |
| Q9H4F8 | SMOC1 | SPARC-related modular calcium-binding protein 1 | SPARC-related modular calcium-binding protein 1 | SMOC1 | SMOC1 | 100% | 100% |
| P60880 | SNAP25 | Synaptosome Associated Protein 25 | Synaptosomal-associated protein 25 | SNAP25 | SNAP25 | 100% | 100% |
| P37840 | SNCA | Synuclein Alpha | Alpha-synuclein | α-Syn | SNCA | 98% | 100% |
| Q16143 | SNCB | Beta-synuclein | Beta-synuclein | β-syn | SNCB | 100% | 100% |
| P00441 | SOD1 | Superoxide Dismutase 1 | Superoxide dismutase [Cu-Zn] | SOD1 | SOD1 | 100% | 100% |
| Q13501 | SQSTM1 | Sequestosome 1 | Sequestosome-1 | SQSTM1 | SQSTM1 | 100% | 100% |
| Q7Z5A7 | TAFA5 | TAFA Chemokine Family Member 5 | Chemokine-like protein TAFA-5 | TAFA5 | TAFA5 | 100% | 100% |
| Q13148 | TARDBP | TAR DNA Binding Protein | TAR DNA-binding protein 43 | TDP43 | TARDBP | 90% | 100% |
| Q02763 | TEK | TEK Receptor Tyrosine Kinase | Angiopoietin-1 receptor | Tie-2/TEK | TEK | 83% | 100% |
| P35625 | TIMP3 | TIMP Metallopeptidase Inhibitor 3 | Metalloproteinase inhibitor 3 | TIMP3 | TIMP3 | 100% | 100% |
| P01375 | TNF | Tumor Necrosis Factor | Tumor necrosis factor | TNF-a | TNF | 100% | 100% |
| Q9NP99 | TREM1 | Triggering Receptor Expressed On Myeloid Cells 1 | Triggering Receptor Expressed On Myeloid Cells 2 | sTREM1 | TREM1 | 91% | 100% |
| Q9NZC2 | TREM2 | Triggering Receptor Expressed on Myeloid Cells 2 | Triggering receptor expressed on myeloid cells 2 | TREM2 | TREM2 | 100% | 100% |
| P0CG47 | UBB | Polyubiquitin-B | Polyubiquitin-B | UBB | UBB | 100% | 100% |
| P09936 | UCHL1 | Ubiquitin C-Terminal Hydrolase L1 | Ubiquitin C-terminal hydrolase L1 | UCHL1 | UCHL1 | 100% | 100% |
| P19320 | VCAM1 | Vascular Cell Adhesion Molecule 1 | Vascular cell adhesion protein 1 | VCAM1/CD106 | VCAM1 | 100% | 100% |
| P15692 | VEGFA | Vascular Endothelial Growth Factor A | Vascular endothelial growth factor A | VEGF-A | VEGFA | 100% | 100% |
| O43915 | VEGFD | Vascular Endothelial Growth Factor D | Vascular endothelial growth factor D | VEGF-D | VEGFD | 100% | 100% |
| O15240 | VGF | Neurosecretory protein VGF | Neurosecretory protein VGF | VGF | VGF | 100% | 100% |
| P62760 | VSNL1 | Visinin Like 1 | Visinin-like protein 1 | VILIP-1 | VSNL1 | 100% | 100% |
| P61981 | YWHAG | Tyrosine 3-Monooxygenase/Tryptophan 5-Monooxygenase Activation Protein Gamma | 14-3-3 protein gamma-B | YWHAG | YWHAG | 100% | 100% |
| P63104 | YWHAZ | Tyrosine 3-Monooxygenase/Tryptophan 5-Monooxygenase Activation Protein Zeta | 14-3-3 protein zeta/delta | YWHAZ | YWHAZ | 100% | 100% |

**Legend**: The table provides UniProt IDs, gene names, full protein names, and common abbreviations for each biomarker. The proportion of subjects with detectable signal in CSF and plasma is shown. * only E4 carrier were accounted.

**Supplementary Table 2:** Correlation between CSF and plasma NULISA biomarker concentrations.

| **Biomarker** | **rho** | **P value** |
| --- | --- | --- |
| Ab38 | 0.141 | 0.0653 |
| Ab40 | 0.144 | 0.0612 |
| Ab42 | 0.245 | 0.0013 |
| Ab4240 | 0.272 | 0.0003 |
| ACHE | 0.072 | 0.3505 |
| AGRN | 0.147 | 0.0557 |
| ANXA5 | 0.103 | 0.1894 |
| APOE | -0.031 | 0.6893 |
| APOE4* | 0.271 | 0.0281 |
| ARSA | 0.095 | 0.416 |
| BACE1 | 0.116 | 0.131 |
| BASP1 | 0.226 | 0.003 |
| CALB2 | 0.112 | 0.146 |
| CCL11 | 0.230 | 0.0024 |
| CCL13 | 0.165 | 0.1156 |
| CCL2 | 0.135 | 0.0791 |
| CCL22 | 0.204 | 0.0907 |
| CCL26 | 0.256 | 0.0008 |
| CCL3 | 0.361 | <0.0001 |
| CCL4 | 0.373 | <0.0001 |
| CD63 | 0.039 | 0.6115 |
| CHI3L1 | 0.169 | 0.0272 |
| CHIT1 | 0.543 | <0.0001 |
| CNTN2 | 0.049 | 0.5501 |
| CRH | -0.023 | 0.7613 |
| CRP | 0.856 | <0.0001 |
| CSF2 | 0.222 | 0.0036 |
| CST3 | 0.000 | 0.9978 |
| CX3CL1 | 0.120 | 0.1179 |
| CXCL1 | 0.131 | 0.089 |
| CXCL10 | 0.221 | 0.0037 |
| CXCL8 | 0.106 | 0.1692 |
| DDC | 0.119 | 0.1685 |
| ENO2 | 0.019 | 0.808 |
| FABP3 | 0.182 | 0.0171 |
| FGF2 | 0.021 | 0.7881 |
| FLT1 | 0.125 | 0.1042 |
| FOLR1 | -0.085 | 0.2665 |
| GDF15 | 0.703 | <0.0001 |
| GDI1 | -0.012 | 0.88 |
| GDNF | 0.113 | 0.1395 |
| GFAP | 0.428 | <0.0001 |
| GOT1 | 0.156 | 0.042 |
| HTT | -0.046 | 0.5518 |
| ICAM1 | 0.129 | 0.2678 |
| IFNG | 0.274 | 0.0003 |
| IGF1R | 0.146 | 0.0564 |
| IGFBP7 | 0.144 | 0.0599 |
| IL10 | 0.245 | 0.0013 |
| IL12p70 | 0.172 | 0.0243 |
| IL13 | 0.436 | <0.0001 |
| IL15 | 0.179 | 0.0193 |
| IL16 | 0.141 | 0.0665 |
| IL17A | -0.014 | 0.8567 |
| IL18 | 0.378 | <0.0001 |
| IL1B | 0.142 | 0.0639 |
| IL2 | 0.186 | 0.0205 |
| IL33 | 0.043 | 0.579 |
| IL4 | 0.175 | 0.0218 |
| IL5 | 0.156 | 0.0421 |
| IL6 | 0.247 | 0.0012 |
| IL6R | 0.230 | 0.0045 |
| IL7 | -0.033 | 0.6639 |
| IL9 | 0.042 | 0.586 |
| KDR | 0.549 | <0.0001 |
| KLK6 | -0.052 | 0.503 |
| MAPT | 0.407 | <0.0001 |
| MDH1 | 0.091 | 0.2366 |
| MSLN | 0.346 | <0.0001 |
| NEFH | 0.238 | 0.0018 |
| NEFL | 0.497 | <0.0001 |
| NGF | 0.014 | 0.896 |
| NPTX1 | -0.079 | 0.3018 |
| NPTX2 | -0.034 | 0.6631 |
| NPTXR | 0.135 | 0.0778 |
| NPY | 0.221 | 0.0037 |
| NRGN | -0.007 | 0.9296 |
| Oligo_SNCA | 0.056 | 0.4645 |
| PARK7 | 0.057 | 0.4619 |
| PDGFRB | 0.724 | <0.0001 |
| PDLIM5 | 0.006 | 0.9433 |
| PGF | 0.230 | 0.0026 |
| PRDX6 | 0.092 | 0.2334 |
| PSEN1 | -0.148 | 0.0541 |
| pSNCA_129 | 0.081 | 0.2939 |
| pTau_181 | 0.666 | <0.0001 |
| pTau_217 | 0.793 | <0.0001 |
| pTau_231 | 0.713 | <0.0001 |
| pTDP43_409 | 0.114 | 0.1389 |
| REST | -0.008 | 0.9187 |
| RUVBL2 | 0.028 | 0.7178 |
| S100B | 0.269 | 0.0004 |
| SFRP1 | 0.054 | 0.4842 |
| SFTPD | 0.329 | <0.0001 |
| SLIT2 | 0.115 | 0.1338 |
| SMOC1 | 0.024 | 0.7544 |
| SNAP25 | -0.083 | 0.2798 |
| SNCA | 0.102 | 0.1917 |
| SNCB | 0.146 | 0.0573 |
| SOD1 | 0.090 | 0.2399 |
| SQSTM1 | 0.125 | 0.104 |
| TAFA5 | -0.029 | 0.7078 |
| TARDBP | 0.011 | 0.8955 |
| TEK | 0.053 | 0.5341 |
| TIMP3 | 0.034 | 0.6619 |
| TNF | 0.229 | 0.0026 |
| TREM1 | 0.131 | 0.1055 |
| TREM2 | 0.292 | 0.0001 |
| UBB | 0.266 | 0.0005 |
| UCHL1 | 0.073 | 0.34 |
| VCAM1 | 0.270 | 0.0004 |
| VEGFA | 0.004 | 0.9564 |
| VEGFD | 0.152 | 0.0471 |
| VGF | 0.064 | 0.4056 |
| VSNL1 | 0.244 | 0.0014 |
| YWHAG | -0.061 | 0.4243 |
| YWHAZ | -0.127 | 0.0971 |

**Legend**: Spearman’s rank correlation coefficients (rho) are shown together with corresponding p-values. * only E4 carrier were accounted.

**Supplementary Table 3:** Association between NULISA CSF and plasma biomarker concentrations, with amyloid status and MCI conversion

|  |  | **Converters vs non converters** | | **Amyloid positive vs negative** | |
| --- | --- | --- | --- | --- | --- |
| **CSF Biomarker** | **N** | **P vlaue (age. sex. E4)** | **P vlaue (age. sex. E4) FDR** | **P vlaue (age. sex. E4)** | **P vlaue (age. sex. E4) FDR** |
| ACHE | 171 | 0.6795 | 0.9324 | 0.1601 | 0.3778 |
| AGRN | 171 | 0.2759 | 0.7542 | 0.2449 | 0.4737 |
| ANXA5 | 164 | 0.9453 | 0.9815 | 0.8149 | 0.8903 |
| APOE | 171 | 0.6275 | 0.9324 | 0.2047 | 0.4391 |
| APOE4 | 171 | 0.0392 | 0.2648 | 0.3167 | 0.5543 |
| ARSA | 76 | 0.9511 | 0.9815 | 0.5176 | 0.7307 |
| Ab38 | 171 | 0.4413 | 0.8265 | 0.8139 | 0.8903 |
| Ab40 | 171 | 0.0994 | 0.4189 | 0.6978 | 0.8724 |
| Ab42 | 171 | <0.0001 | 0.0009 | <0.0001 | <0.0001 |
| Ab4240 | 171 | 0.0001 | 0.0014 | <0.0001 | <0.0001 |
| BACE1 | 171 | 0.9972 | 0.9972 | 0.0684 | 0.2604 |
| BASP1 | 171 | 0.7871 | 0.9657 | 0.7445 | 0.8786 |
| CALB2 | 171 | 0.2822 | 0.7542 | 0.5384 | 0.7387 |
| CCL11 | 171 | 0.5712 | 0.9324 | 0.4428 | 0.6876 |
| CCL13 | 92 | 0.6247 | 0.9324 | 0.7171 | 0.8724 |
| CCL2 | 171 | 0.2337 | 0.6727 | 0.7268 | 0.8752 |
| CCL22 | 70 | 0.158 | 0.526 | 0.4992 | 0.7272 |
| CCL26 | 171 | 0.3837 | 0.7675 | 0.4973 | 0.7272 |
| CCL3 | 171 | 0.3703 | 0.7675 | 0.2093 | 0.441 |
| CCL4 | 168 | 0.1605 | 0.526 | 0.1543 | 0.3716 |
| CD63 | 171 | 0.8984 | 0.9815 | 0.0224 | 0.1261 |
| CHI3L1 | 171 | 0.6654 | 0.9324 | 0.0375 | 0.1526 |
| CHIT1 | 167 | 0.9084 | 0.9815 | <0.0001 | 0.0001 |
| CNTN2 | 171 | 0.6795 | 0.9324 | 0.7132 | 0.8724 |
| CRH | 171 | 0.9639 | 0.9815 | 0.8654 | 0.92 |
| CRP | 171 | 0.7174 | 0.9465 | 0.1285 | 0.3525 |
| CSF2 | 171 | 0.1133 | 0.461 | 0.71 | 0.8724 |
| CST3 | 171 | 0.0876 | 0.4018 | 0.5509 | 0.7472 |
| CX3CL1 | 171 | 0.3258 | 0.7542 | 0.4509 | 0.6909 |
| CXCL1 | 171 | 0.335 | 0.7542 | 0.0604 | 0.2377 |
| CXCL10 | 171 | 0.0157 | 0.1428 | 0.4146 | 0.6703 |
| CXCL8 | 171 | 0.2162 | 0.6468 | 0.7399 | 0.8786 |
| DDC | 136 | 0.9702 | 0.9815 | 0.8642 | 0.92 |
| ENO2 | 171 | 0.3452 | 0.7542 | 0.0177 | 0.1042 |
| FABP3 | 171 | 0.1713 | 0.5464 | <0.0001 | 0.0002 |
| FGF2 | 171 | 0.6041 | 0.9324 | 0.2334 | 0.4737 |
| FLT1 | 171 | 0.7299 | 0.9465 | 0.0907 | 0.2974 |
| FOLR1 | 171 | 0.3346 | 0.7542 | 0.1501 | 0.3691 |
| GDF15 | 160 | 0.888 | 0.9815 | 0.1343 | 0.3601 |
| GDI1 | 171 | 0.0015 | 0.0201 | 0.0261 | 0.1341 |
| GDNF | 171 | 0.1247 | 0.4905 | 0.5355 | 0.7387 |
| GFAP | 171 | 0.0799 | 0.4018 | 0.002 | 0.0158 |
| GOT1 | 171 | 0.9223 | 0.9815 | 0.0327 | 0.1486 |
| HTT | 171 | 0.5858 | 0.9324 | 0.1264 | 0.3525 |
| ICAM1 | 75 | 0.2066 | 0.6416 | 0.037 | 0.1526 |
| IFNG | 171 | 0.3111 | 0.7542 | 0.5751 | 0.7712 |
| IGF1R | 171 | 0.5222 | 0.8959 | 0.2883 | 0.5339 |
| IGFBP7 | 171 | 0.3812 | 0.7675 | 0.1174 | 0.3378 |
| IL10 | 171 | 0.9692 | 0.9815 | 0.5202 | 0.7307 |
| IL12p70 | 171 | 0.465 | 0.8441 | 0.8052 | 0.8903 |
| IL13 | 171 | 0.943 | 0.9815 | 0.1085 | 0.3283 |
| IL15 | 171 | 0.4063 | 0.7859 | 0.0321 | 0.1486 |
| IL16 | 171 | 0.0919 | 0.4018 | 0.1744 | 0.3958 |
| IL17A | 171 | 0.6249 | 0.9324 | 0.2426 | 0.4737 |
| IL18 | 146 | 0.3139 | 0.7542 | 0.0861 | 0.2923 |
| IL1B | 171 | 0.7225 | 0.9465 | 0.4649 | 0.6978 |
| IL2 | 156 | 0.0633 | 0.3556 | 0.6696 | 0.8652 |
| IL33 | 171 | 0.5105 | 0.8959 | 0.357 | 0.5933 |
| IL4 | 171 | 0.2193 | 0.6468 | 0.7815 | 0.8889 |
| IL5 | 171 | 0.9273 | 0.9815 | 0.698 | 0.8724 |
| IL6 | 171 | 0.8119 | 0.9657 | 0.294 | 0.5339 |
| IL6R | 152 | 0.711 | 0.9465 | 0.9045 | 0.9362 |
| IL7 | 171 | 0.0905 | 0.4018 | 0.8031 | 0.8903 |
| IL9 | 171 | 0.5835 | 0.9324 | 0.2941 | 0.5339 |
| KDR | 157 | 0.5239 | 0.8959 | 0.7708 | 0.8889 |
| KLK6 | 171 | 0.7459 | 0.9465 | 0.1411 | 0.3619 |
| MAPT | 171 | <0.0001 | 0.0002 | <0.0001 | <0.0001 |
| MDH1 | 171 | 0.3134 | 0.7542 | 0.0008 | 0.007 |
| MSLN | 171 | 0.74 | 0.9465 | 0.9489 | 0.9652 |
| NEFH | 171 | 0.0369 | 0.2648 | 0.0239 | 0.1284 |
| NEFL | 171 | 0.0002 | 0.0031 | 0.0027 | 0.02 |
| NGF | 89 | 0.1316 | 0.5008 | 0.841 | 0.9104 |
| NPTX1 | 171 | 0.599 | 0.9324 | 0.9172 | 0.9411 |
| NPTX2 | 171 | 0.0142 | 0.1428 | 0.6746 | 0.8652 |
| NPTXR | 171 | 0.0824 | 0.4018 | 0.9682 | 0.9682 |
| NPY | 171 | 0.0529 | 0.3283 | 0.9663 | 0.9682 |
| NRGN | 171 | 0.0157 | 0.1428 | 0.0005 | 0.0052 |
| Oligo_SNCA | 171 | 0.8125 | 0.9657 | 0.2874 | 0.5339 |
| PARK7 | 171 | 0.8042 | 0.9657 | 0.5831 | 0.7731 |
| PDGFRB | 167 | 0.8983 | 0.9815 | 0.4247 | 0.6772 |
| PDLIM5 | 171 | 0.0404 | 0.2648 | 0.5133 | 0.7307 |
| PGF | 171 | 0.7737 | 0.961 | 0.7595 | 0.8873 |
| PRDX6 | 171 | 0.6461 | 0.9324 | 0.0867 | 0.2923 |
| PSEN1 | 171 | 0.6455 | 0.9324 | 0.1487 | 0.3691 |
| pSNCA_129 | 171 | 0.3039 | 0.7542 | 0.0751 | 0.276 |
| pTau_181 | 171 | <0.0001 | 0.0002 | <0.0001 | <0.0001 |
| pTau_217 | 171 | <0.0001 | 0.0002 | <0.0001 | <0.0001 |
| pTau_231 | 171 | <0.0001 | 0.0002 | <0.0001 | <0.0001 |
| pTDP43_409 | 171 | 0.087 | 0.4018 | 0.8841 | 0.9314 |
| PTN | 171 | 0.4038 | 0.7859 | 0.3197 | 0.5543 |
| REST | 171 | 0.1462 | 0.5199 | 0.3241 | 0.5543 |
| RUVBL2 | 171 | 0.4388 | 0.8265 | 0.2366 | 0.4737 |
| S100B | 171 | 0.8184 | 0.9657 | 0.0048 | 0.0321 |
| SFRP1 | 171 | 0.4591 | 0.8441 | 0.1974 | 0.4313 |
| SFTPD | 169 | 0.3404 | 0.7542 | 0.654 | 0.8574 |
| SLIT2 | 171 | 0.3235 | 0.7542 | 0.0123 | 0.0766 |
| SMOC1 | 171 | 0.056 | 0.3304 | <0.0001 | <0.0001 |
| SNAP25 | 171 | 0.3595 | 0.7675 | 0.1126 | 0.332 |
| SNCA | 167 | 0.6627 | 0.9324 | 0.0049 | 0.0321 |
| SNCB | 171 | 0.0099 | 0.1167 | 0.0002 | 0.0018 |
| SOD1 | 171 | 0.9141 | 0.9815 | 0.0375 | 0.1526 |
| SQSTM1 | 171 | 0.2666 | 0.749 | 0.2371 | 0.4737 |
| TAFA5 | 171 | 0.9732 | 0.9815 | 0.3326 | 0.5607 |
| TARDBP | 153 | 0.7332 | 0.9465 | 0.1007 | 0.3192 |
| TEK | 141 | 0.9674 | 0.9815 | 0.3133 | 0.5543 |
| TIMP3 | 171 | 0.5746 | 0.9324 | 0.0772 | 0.276 |
| TNF | 171 | 0.5156 | 0.8959 | 0.4671 | 0.6978 |
| TREM1 | 155 | 0.0275 | 0.2161 | 0.0314 | 0.1486 |
| TREM2 | 171 | 0.7556 | 0.9485 | 0.4133 | 0.6703 |
| UBB | 171 | 0.0242 | 0.2036 | 0.1385 | 0.3619 |
| UCHL1 | 171 | <0.0001 | 0.0005 | <0.0001 | 0.0001 |
| VCAM1 | 171 | 0.5654 | 0.9324 | 0.1028 | 0.3192 |
| VEGFA | 171 | 0.6561 | 0.9324 | 0.1805 | 0.4018 |
| VEGFD | 171 | 0.9202 | 0.9815 | 0.4384 | 0.6876 |
| VGF | 171 | 0.379 | 0.7675 | 0.9044 | 0.9362 |
| VSNL1 | 171 | 0.1498 | 0.5199 | 0.001 | 0.0085 |
| YWHAG | 171 | 0.1393 | 0.5138 | 0.1739 | 0.3958 |
| YWHAZ | 171 | 0.935 | 0.9815 | 0.7835 | 0.8889 |

|  |  | **Converters vs non converters** | | **Amyloid positive vs negative** | |
| --- | --- | --- | --- | --- | --- |
| **Plsama Biomarker** | **N** | **P vlaue (age. sex. E4)** | **P vlaue (age. sex. E4) FDR** | **P vlaue (age. sex. E4)** | **P vlaue (age. sex. E4) FDR** |
| Ab38 | 173 | 0.4921 | 0.8054 | 0.6418 | 0.8006 |
| Ab40 | 173 | 0.6571 | 0.9225 | 0.643 | 0.8006 |
| Ab42 | 173 | 0.4277 | 0.7939 | 0.0063 | 0.0778 |
| Ab4240 | 173 | 0.7243 | 0.9303 | 0.0012 | 0.0259 |
| ACHE | 173 | 0.1438 | 0.6365 | 0.0592 | 0.2595 |
| AGRN | 173 | 0.7179 | 0.9303 | 0.7444 | 0.8517 |
| ANXA5 | 173 | 0.1323 | 0.6224 | 0.0615 | 0.2605 |
| APOE | 173 | 0.3221 | 0.7504 | 0.8748 | 0.9336 |
| APOE4 | 173 | 0.0583 | 0.4973 | 0.2215 | 0.5252 |
| ARSA | 173 | 0.259 | 0.7401 | 0.0128 | 0.1086 |
| BACE1 | 173 | 0.5837 | 0.8636 | 0.0412 | 0.2179 |
| BASP1 | 173 | 0.4533 | 0.7997 | 0.0074 | 0.0778 |
| BDNF | 173 | 0.7115 | 0.9303 | 0.0698 | 0.2769 |
| CALB2 | 173 | 0.8948 | 0.9502 | 0.3215 | 0.6005 |
| CCL11 | 173 | 0.1654 | 0.6365 | 0.3108 | 0.5981 |
| CCL13 | 173 | 0.3585 | 0.7646 | 0.0209 | 0.1541 |
| CCL17 | 173 | 0.4481 | 0.7997 | 0.6835 | 0.8181 |
| CCL2 | 173 | 0.3742 | 0.7664 | 0.5979 | 0.7828 |
| CCL22 | 173 | 0.3368 | 0.7504 | 0.9732 | 0.9968 |
| CCL26 | 173 | 0.4946 | 0.8054 | 0.2055 | 0.5252 |
| CCL3 | 173 | 0.7786 | 0.9303 | 0.6059 | 0.7852 |
| CCL4 | 173 | 0.5754 | 0.8636 | 0.8905 | 0.9425 |
| CD40LG | 173 | 0.7227 | 0.9303 | 0.0693 | 0.2769 |
| CD63 | 173 | 0.4201 | 0.7939 | 0.783 | 0.8801 |
| CHI3L1 | 173 | 0.8675 | 0.9429 | 0.0491 | 0.2399 |
| CHIT1 | 169 | 0.3627 | 0.7646 | 0.5849 | 0.7773 |
| CNTN2 | 150 | 0.332 | 0.7504 | 0.0742 | 0.2773 |
| CRH | 173 | 0.6813 | 0.9303 | 0.3196 | 0.6005 |
| CRP | 173 | 0.4921 | 0.8054 | 0.0218 | 0.1541 |
| CSF2 | 173 | 0.6772 | 0.9303 | 0.7003 | 0.8235 |
| CST3 | 173 | 0.8761 | 0.9429 | 0.5074 | 0.724 |
| CX3CL1 | 173 | 0.1967 | 0.6573 | 0.3058 | 0.5975 |
| CXCL1 | 173 | 0.5667 | 0.8636 | 0.5875 | 0.7773 |
| CXCL10 | 173 | 0.1271 | 0.6224 | 0.8383 | 0.9114 |
| CXCL8 | 173 | 0.3672 | 0.7646 | 0.8512 | 0.9161 |
| DDC | 173 | 0.7597 | 0.9303 | 0.6327 | 0.8006 |
| ENO2 | 173 | 0.7537 | 0.9303 | 0.408 | 0.6642 |
| FABP3 | 173 | 0.5979 | 0.8636 | 0.4931 | 0.7165 |
| FCN2 | 173 | 0.5501 | 0.8636 | 0.3641 | 0.6291 |
| FGF2 | 173 | 0.0212 | 0.3372 | 0.128 | 0.393 |
| FLT1 | 173 | 0.8621 | 0.9429 | 0.2048 | 0.5252 |
| FOLR1 | 173 | 0.1281 | 0.6224 | 0.0342 | 0.2068 |
| GDF15 | 162 | 0.8083 | 0.9303 | 0.4569 | 0.7165 |
| GDI1 | 173 | 0.0498 | 0.4973 | 0.1362 | 0.393 |
| GDNF | 173 | 0.4804 | 0.8054 | 0.3899 | 0.6431 |
| GFAP | 173 | 0.0523 | 0.4973 | <0.0001 | 0.0001 |
| GOT1 | 173 | 0.2914 | 0.7504 | 0.0813 | 0.2868 |
| HBA1 | 132 | 0.0862 | 0.6224 | 0.0998 | 0.327 |
| HTT | 173 | 0.1036 | 0.6224 | 0.038 | 0.2119 |
| ICAM1 | 173 | 0.8992 | 0.9502 | 0.0261 | 0.1654 |
| IFNG | 173 | 0.8657 | 0.9429 | 0.2999 | 0.5952 |
| IGF1R | 173 | 0.9544 | 0.9775 | 0.3515 | 0.6243 |
| IGFBP7 | 173 | 0.8694 | 0.9429 | 0.253 | 0.5356 |
| IL10 | 173 | 0.9736 | 0.9847 | 0.6826 | 0.8181 |
| IL12p70 | 173 | 0.2292 | 0.7401 | 0.146 | 0.412 |
| IL13 | 173 | 0.4886 | 0.8054 | 0.1533 | 0.4233 |
| IL15 | 173 | 0.1156 | 0.6224 | 0.5608 | 0.7636 |
| IL16 | 173 | 0.2576 | 0.7401 | 0.4946 | 0.7165 |
| IL17A | 173 | 0.5984 | 0.8636 | 0.661 | 0.8072 |
| IL18 | 173 | 0.9923 | 0.9923 | 0.7769 | 0.8801 |
| IL1B | 173 | 0.254 | 0.7401 | 0.231 | 0.5252 |
| IL2 | 173 | 0.7437 | 0.9303 | 0.4706 | 0.7165 |
| IL33 | 173 | 0.769 | 0.9303 | 0.7988 | 0.8898 |
| IL4 | 173 | 0.098 | 0.6224 | 0.227 | 0.5252 |
| IL5 | 173 | 0.661 | 0.9225 | 0.3539 | 0.6243 |
| IL6 | 173 | 0.4072 | 0.7939 | 0.2869 | 0.5821 |
| IL6R | 173 | 0.1518 | 0.6365 | 0.9497 | 0.9968 |
| IL7 | 173 | 0.6097 | 0.87 | 0.052 | 0.2447 |
| IL9 | 173 | 0.2645 | 0.7401 | 0.4759 | 0.7165 |
| KDR | 170 | 0.4423 | 0.7997 | 0.3399 | 0.6167 |
| KLK6 | 173 | 0.9224 | 0.9524 | 0.6893 | 0.8181 |
| MAPT | 173 | 0.0134 | 0.3335 | <0.0001 | 0.0012 |
| MDH1 | 173 | 0.5946 | 0.8636 | 0.0039 | 0.0667 |
| MME | 173 | 0.087 | 0.6224 | 0.3715 | 0.6291 |
| MSLN | 173 | 0.8128 | 0.9303 | 0.7215 | 0.8407 |
| NEFH | 173 | 0.1319 | 0.6224 | 0.0384 | 0.2119 |
| NEFL | 173 | 0.1761 | 0.6404 | 0.1123 | 0.3564 |
| NGF | 173 | 0.1906 | 0.6541 | 0.4213 | 0.6773 |
| NPTX1 | 173 | 0.5964 | 0.8636 | 0.8252 | 0.9113 |
| NPTX2 | 173 | 0.3124 | 0.7504 | 0.9897 | 0.9975 |
| NPTXR | 173 | 0.7355 | 0.9303 | 0.9579 | 0.9968 |
| NPY | 173 | 0.8218 | 0.9319 | 0.2214 | 0.5252 |
| NRGN | 173 | 0.0315 | 0.445 | 0.2171 | 0.5252 |
| Oligo_SNCA | 173 | 0.4313 | 0.7939 | 0.0195 | 0.1541 |
| PARK7 | 173 | 0.3007 | 0.7504 | 0.0244 | 0.1634 |
| PDGFRB | 173 | 0.4012 | 0.7939 | 0.3303 | 0.608 |
| PDLIM5 | 173 | 0.8679 | 0.9429 | 0.2379 | 0.5252 |
| PGF | 173 | 0.1792 | 0.6404 | 0.0794 | 0.2868 |
| PGK1 | 173 | 0.3096 | 0.7504 | 0.0126 | 0.1086 |
| POSTN | 173 | 0.0986 | 0.6224 | 0.1312 | 0.393 |
| PRDX6 | 173 | 0.7655 | 0.9303 | 0.007 | 0.0778 |
| PSEN1 | 173 | 0.8094 | 0.9303 | 0.5652 | 0.7636 |
| pSNCA_129 | 173 | 0.2393 | 0.7401 | 0.011 | 0.1073 |
| pTau_181 | 173 | 0.0002 | 0.0096 | <0.0001 | <0.0001 |
| pTau_217 | 173 | <0.0001 | 0.002 | <0.0001 | <0.0001 |
| pTau_231 | 173 | 0.0003 | 0.0123 | <0.0001 | <0.0001 |
| pTDP43_409 | 173 | 0.0158 | 0.3335 | 0.176 | 0.4755 |
| REST | 173 | 0.1571 | 0.6365 | 0.3822 | 0.6387 |
| RUVBL2 | 173 | 0.0471 | 0.4973 | 0.0737 | 0.2773 |
| S100A12 | 173 | 0.0555 | 0.4973 | 0.5346 | 0.7517 |
| S100B | 173 | 0.5895 | 0.8636 | 0.5386 | 0.7517 |
| SAA1 | 173 | 0.9054 | 0.9502 | 0.0462 | 0.2348 |
| SFRP1 | 173 | 0.1604 | 0.6365 | 0.4879 | 0.7165 |
| SFTPD | 173 | 0.721 | 0.9303 | 0.5459 | 0.7536 |
| SLIT2 | 173 | 0.9181 | 0.9524 | 0.4771 | 0.7165 |
| SMOC1 | 173 | 0.1565 | 0.6365 | 0.6191 | 0.7942 |
| SNAP25 | 173 | 0.2644 | 0.7401 | 0.736 | 0.8498 |
| SNCA | 173 | 0.4281 | 0.7939 | 0.0052 | 0.0728 |
| SNCB | 173 | 0.2681 | 0.7401 | 0.9964 | 0.9975 |
| SOD1 | 173 | 0.1815 | 0.6404 | 0.0042 | 0.0667 |
| SQSTM1 | 173 | 0.4306 | 0.7939 | 0.8397 | 0.9114 |
| TAFA5 | 173 | 0.4773 | 0.8054 | 0.4833 | 0.7165 |
| TARDBP | 173 | 0.0208 | 0.3372 | 0.0565 | 0.2562 |
| TEK | 173 | 0.321 | 0.7504 | 0.4965 | 0.7165 |
| TIMP3 | 173 | 0.3516 | 0.7646 | 0.1004 | 0.327 |
| TNF | 173 | 0.8044 | 0.9303 | 0.263 | 0.5476 |
| TREM1 | 173 | 0.5675 | 0.8636 | 0.9975 | 0.9975 |
| TREM2 | 173 | 0.9769 | 0.9847 | 0.1361 | 0.393 |
| UBB | 173 | 0.1142 | 0.6224 | 0.2362 | 0.5252 |
| UCHL1 | 173 | 0.3173 | 0.7504 | 0.6552 | 0.8072 |
| VCAM1 | 173 | 0.0587 | 0.4973 | 0.2175 | 0.5252 |
| VEGFA | 173 | 0.8092 | 0.9303 | 0.9679 | 0.9968 |
| VEGFD | 173 | 0.1123 | 0.6224 | 0.0904 | 0.3103 |
| VGF | 173 | 0.8131 | 0.9303 | 0.2887 | 0.5821 |
| VSNL1 | 173 | 0.3115 | 0.7504 | 0.244 | 0.5252 |
| YWHAG | 173 | 0.0006 | 0.0181 | 0.2421 | 0.5252 |
| YWHAZ | 173 | 0.3291 | 0.7504 | 0.3679 | 0.6291 |

**Legend**: The table shows p-values from Cox regression models adjusted for age, sex, and APOE4 genotype. The false discovery rate (FDR) correction was applied for multiple comparisons.

**Supplementary Table 4:** **Performance of plasma biomarkers and their combinations for prediction of MCI conversion**.

| **Num** | **CSF Biomarker** | **AUC** | **CI_lower** | **CI_upper** | **Regression coefficients** | **Log Rank P value** | **HR 1st vs 3rd** |
| --- | --- | --- | --- | --- | --- | --- | --- |
| 3 | IL16 - MAPT - NPTX2 | 0.86 | 0.80 | 0.91 | 8.9503 -1.6527*IL16 + 1.9436*MAPT -1.7068*NPTX2 | 7.83E-17 | 39.8 (9.6-165.2) |
| 3 | NPTX2 - PDLIM5 - pTau_181 | 0.83 | 0.77 | 0.89 | -16.3979 -1.4083*NPTX2 + 1.9152*PDLIM5 + 1.2287*pTau_181 | 1.16E-12 | 17.3 (6.1-48.8) |
| 3 | IL16 - NPTX2 - pTau_181 | 0.83 | 0.77 | 0.89 | 4.2311 -1.2308*IL16 -1.2185*NPTX2 + 1.3108*pTau_181 | 1.76E-10 | 15 (5.3-42.3) |
| 3 | CXCL10 - NPTX2 - pTau_181 | 0.83 | 0.76 | 0.88 | 4.5865 -0.6862*CXCL10 -1.2935*NPTX2 + 1.2051*pTau_181 | 9.15E-13 | 21.6 (6.6-70.4) |
| 3 | NEFL - NPTX2 - pTau_181 | 0.83 | 0.76 | 0.89 | -10.9475 + 0.4997*NEFL -1.175*NPTX2 + 1.0575*pTau_181 | 1.55E-12 | 13.9 (5.4-35.7) |
| 2 | MAPT - NPTX2 | 0.83 | 0.76 | 0.88 | -0.1981 + 1.5609*MAPT -1.6494*NPTX2 | 8.66E-11 | 15.8 (5.6-44.8) |
| 3 | MAPT - NPTX2 - pTau_181 | 0.82 | 0.76 | 0.88 | -0.0926 + 1.6076*MAPT -1.6586*NPTX2 -0.038*pTau_181 | 8.66E-11 | 15.8 (5.6-44.8) |
| 2 | NPTX2 - pTau_181 | 0.81 | 0.75 | 0.87 | -2.6105 -1.2526*NPTX2 + 1.1334*pTau_181 | 5.18E-13 | 13.7 (5.3-35.2) |
| 3 | IL16 - PDLIM5 - pTau_181 | 0.81 | 0.73 | 0.87 | -22.8706 -1.5084*IL16 + 2.0406*PDLIM5 + 1.1134*pTau_181 | 6.11E-09 | 8.1 (3.6-18.2) |
| 3 | CXCL10 - IL16 - pTau_181 | 0.79 | 0.72 | 0.86 | -3.1941 -0.5733*CXCL10 -1.1219*IL16 + 1.0504*pTau_181 | 1.37E-09 | 7.1 (3.3-15.4) |
| 2 | IL16 - pTau_181 | 0.77 | 0.70 | 0.84 | -7.9988 -1.2926*IL16 + 1.0331*pTau_181 | 7.51E-10 | 6.4 (3.1-13.4) |
| 2 | PDLIM5 - pTau_181 | 0.77 | 0.69 | 0.85 | -27.5547 + 1.4802*PDLIM5 + 0.8891*pTau_181 | 6.50E-09 | 7.3 (3.4-15.7) |
| 2 | MAPT - pTau_181 | 0.75 | 0.67 | 0.82 | -15.6047 -0.1044*MAPT + 0.9292*pTau_181 | 2.79E-09 | 5.3 (2.7-10.4) |
| 1 | pTau_181 | 0.74 | 0.66 | 0.81 | pTau_181 | 2.07E-09 | 5.9 (2.9-12) |
| 1 | pTau_231 | 0.74 | 0.65 | 0.81 | pTau_231 | 7.07E-09 | 7.3 (3.4-15.9) |
| 1 | MAPT | 0.73 | 0.66 | 0.80 | MAPT | 1.60E-07 | 5.6 (2.8-11.4) |
| 1 | pTau_217 | 0.73 | 0.64 | 0.81 | pTau_217 | 6.19E-09 | 6.5 (3.1-13.6) |
| 1 | Ab42 | 0.72 | 0.63 | 0.80 | Ab42 | 2.73E-05 | 4.8 (2.3-10.2) |
| 1 | UCHL1 | 0.71 | 0.63 | 0.79 | UCHL1 | 2.26E-06 | 5.5 (2.6-11.6) |
| 3 | IL16 - NPTX2 - PDLIM5 | 0.69 | 0.59 | 0.77 | 0.448 -0.4399*IL16 -0.6126*NPTX2 + 1.5601*PDLIM5 | 9.71E-04 | 3.4 (1.7-6.8) |
| 2 | NPTX2 - PDLIM5 | 0.69 | 0.60 | 0.78 | -2.1867 -0.6602*NPTX2 + 1.5026*PDLIM5 | 8.68E-04 | 3.6 (1.8-7.4) |
| 1 | NEFL | 0.68 | 0.59 | 0.77 | NEFL | 4.07E-04 | 3.4 (1.8-6.7) |
| 1 | GDI1 | 0.67 | 0.59 | 0.75 | GDI1 | 5.82E-04 | 3.6 (1.8-7.2) |
| 3 | CXCL10 - IL16 - NPTX2 | 0.65 | 0.57 | 0.73 | 13.8699 -0.4851*CXCL10 -0.118*IL16 -0.5394*NPTX2 | 3.80E-02 | 2.4 (1.2-4.6) |
| 1 | PDLIM5 | 0.65 | 0.56 | 0.74 | PDLIM5 | 2.66E-02 | 2.1 (1.1-4) |
| 2 | IL16 - PDLIM5 | 0.65 | 0.49 | 0.74 | -6.2778 -0.5817*IL16 + 1.4284*PDLIM5 | 9.07E-02 | 2 (1-3.7) |
| 2 | CXCL10 - NPTX2 | 0.65 | 0.55 | 0.73 | 13.2145 -0.4988*CXCL10 -0.5528*NPTX2 | 1.93E-02 | 2.5 (1.3-5) |
| 1 | NRGN | 0.65 | 0.57 | 0.73 | NRGN | 9.46E-04 | 3.2 (1.7-6.2) |
| 2 | IL16 - NPTX2 | 0.63 | 0.50 | 0.72 | 10.0494 -0.3007*IL16 -0.5519*NPTX2 | 2.65E-03 | 3.4 (1.6-6.9) |
| 1 | NPTX2 | 0.63 | 0.53 | 0.72 | NPTX2 | 8.58E-02 | 2 (1-3.7) |
| 2 | CXCL10 - IL16 | 0.60 | 0.45 | 0.68 | 7.3474 -0.5018*CXCL10 -0.2517*IL16 | 2.58E-02 | 1.9 (0.9-3.7) |
| 1 | CXCL10 | 0.58 | 0.44 | 0.67 | CXCL10 | 2.48E-01 | 1.7 (0.9-3.2) |
| 1 | UBB | 0.56 | 0.43 | 0.66 | UBB | 1.35E-01 | 1.8 (0.9-3.5) |
| 1 | SFTPD | 0.55 | 0.47 | 0.63 | SFTPD | 4.23E-01 | 1.3 (0.7-2.4) |
| 1 | IGFBP7 | 0.55 | 0.47 | 0.62 | IGFBP7 | 7.95E-01 | 1.1 (0.6-2.1) |
| 1 | IL16 | 0.54 | 0.46 | 0.63 | IL16 | 9.32E-01 | 1.1 (0.6-2.1) |
| 1 | CCL2 | 0.54 | 0.47 | 0.62 | CCL2 | 6.62E-01 | 1 (0.6-2) |
| 1 | Oligo_SNCA | 0.53 | 0.47 | 0.61 | Oligo_SNCA | 7.91E-01 | 1 (0.5-1.8) |
| 1 | SFRP1 | 0.53 | 0.45 | 0.62 | SFRP1 | 9.53E-01 | 0.9 (0.5-1.6) |
| 1 | FOLR1 | 0.53 | 0.45 | 0.61 | FOLR1 | 8.25E-01 | 0.9 (0.5-1.7) |

| **Num** | **Plasma Biomarker** | **AUC** | **CI_lower** | **CI_upper** | **coefficients** | **Log Rank P value** | **HR 1st vs 3rd** |
| --- | --- | --- | --- | --- | --- | --- | --- |
| 3 | pTau_217 - VCAM1 - YWHAG | 0.81 | 0.75 | 0.87 | 40.7168 + 1.2379*pTau_217 -1.3334*VCAM1 -3.2667*YWHAG | 8.42E-11 | 9.3 (4.1-21) |
| 3 | pTau_217 - pTau_231 - YWHAG | 0.80 | 0.74 | 0.86 | 30.6287 + 1.8949*pTau_217 -0.8995*pTau_231 -3.5168*YWHAG | 2.13E-09 | 7.4 (3.4-16) |
| 3 | APOE4 - pTau_217 - YWHAG | 0.80 | 0.73 | 0.86 | 26.5296 + 0.0422*APOE4 + 1.0757*pTau_217 -3.4554*YWHAG | 4.46E-14 | 8.6 (4.1-17.9) |
| 3 | pTau_181 - pTau_217 - YWHAG | 0.80 | 0.73 | 0.86 | 26.3098 -0.0753*pTau_181 + 1.205*pTau_217 -3.4558*YWHAG | 3.01E-12 | 7.8 (3.8-16.2) |
| 2 | pTau_217 - YWHAG | 0.79 | 0.73 | 0.86 | 25.8546 + 1.1528*pTau_217 -3.454*YWHAG | 7.48E-11 | 7.4 (3.6-15.4) |
| 3 | pTau_181 - VCAM1 - YWHAG | 0.79 | 0.72 | 0.86 | 31.5836 + 1.3852*pTau_181 -1.1641*VCAM1 -3.0917*YWHAG | 3.90E-15 | 9.5 (4.4-20.4) |
| 3 | APOE4 - pTau_181 - YWHAG | 0.79 | 0.72 | 0.86 | 19.8571 + 0.0572*APOE4 + 1.2057*pTau_181 -3.2488*YWHAG | 7.01E-11 | 8 (3.7-17.4) |
| 3 | pTau_181 - pTau_231 - YWHAG | 0.78 | 0.71 | 0.85 | 18.5983 + 1.1262*pTau_181 + 0.16*pTau_231 -3.2123*YWHAG | 6.77E-13 | 7.7 (3.7-16.1) |
| 2 | pTau_181 - YWHAG | 0.78 | 0.70 | 0.85 | 18.4307 + 1.3006*pTau_181 -3.2116*YWHAG | 2.80E-14 | 9.4 (4.4-20.3) |
| 3 | APOE4 - pTau_231 - YWHAG | 0.78 | 0.70 | 0.84 | 21.4641 + 0.0533*APOE4 + 1.0243*pTau_231 -3.1925*YWHAG | 1.26E-09 | 7 (3.4-14.6) |
| 2 | pTau_231 - YWHAG | 0.77 | 0.70 | 0.84 | 20.2133 + 1.1208*pTau_231 -3.1764*YWHAG | 3.07E-10 | 5.6 (2.9-11.1) |
| 3 | APOE4 - pTau_181 - pTau_217 | 0.75 | 0.67 | 0.82 | -11.1388 + 0.0455*APOE4 -0.0093*pTau_181 + 0.9237*pTau_217 | 5.36E-09 | 6.7 (3.2-14) |
| 2 | pTau_217 - pTau_231 | 0.74 | 0.66 | 0.82 | -8.306 + 1.6927*pTau_217 -0.8259*pTau_231 | 1.79E-06 | 5.3 (2.6-10.8) |
| 2 | APOE4 - pTau_217 | 0.74 | 0.66 | 0.81 | -11.1905 + 0.0456*APOE4 + 0.9171*pTau_217 | 5.36E-09 | 6.7 (3.2-14) |
| 2 | APOE4 - pTau_181 | 0.74 | 0.66 | 0.82 | -14.835 + 0.0584*APOE4 + 1.0262*pTau_181 | 1.44E-08 | 5.4 (2.7-10.7) |
| 2 | pTau_181 - pTau_217 | 0.73 | 0.65 | 0.81 | -11.3505 -0.1096*pTau_181 + 1.0836*pTau_217 | 9.25E-08 | 6.5 (3.1-13.5) |
| 2 | pTau_181 - VCAM1 | 0.73 | 0.65 | 0.80 | 0.2258 + 1.2489*pTau_181 -1.3211*VCAM1 | 5.73E-08 | 5.5 (2.7-11.1) |
| 1 | pTau_217 | 0.72 | 0.63 | 0.80 | NA  pTau_217 | 3.98E-07 | 6.1 (2.9-12.8) |
| 1 | pTau_181 | 0.72 | 0.64 | 0.79 | NA  pTau_181 | 1.51E-07 | 5.4 (2.6-10.9) |
| 3 | APOE4 - VCAM1 - YWHAG | 0.72 | 0.64 | 0.79 | 37.2968 + 0.0863*APOE4 -0.7404*VCAM1 -2.4781*YWHAG | 8.60E-05 | 4.2 (2.1-8.5) |
| 1 | pTau_231 | 0.70 | 0.62 | 0.79 | NA  pTau_231 | 5.57E-07 | 5.3 (2.6-10.7) |
| 2 | APOE4 - YWHAG | 0.70 | 0.62 | 0.78 | 28.4703 + 0.0822*APOE4 -2.5765*YWHAG | 2.19E-04 | 3 (1.6-5.6) |
| 2 | VCAM1 - YWHAG | 0.66 | 0.58 | 0.74 | 34.4913 -0.5212*VCAM1 -2.4533*YWHAG | 1.14E-02 | 2.6 (1.3-5.1) |
| 1 | YWHAG | 0.64 | 0.47 | 0.73 | YWHAG | 1.10E-02 | 2.6 (1.3-5.1) |
| 1 | APOE4 | 0.63 | 0.56 | 0.71 | APOE4 | 2.91E-03 | 2.3 (1.4-3.8) |
| 1 | pTDP43_409 | 0.60 | 0.46 | 0.68 | pTDP43_409 | 2.22E-01 | 1.7 (0.9-3.2) |
| 1 | POSTN | 0.59 | 0.49 | 0.67 | POSTN | 2.80E-01 | 1.6 (0.9-3) |
| 1 | S100A12 | 0.59 | 0.49 | 0.67 | S100A12 | 3.17E-02 | 2.4 (1.2-4.6) |
| 1 | IL4 | 0.57 | 0.48 | 0.66 | IL4 | 2.43E-01 | 1.4 (0.7-2.7) |
| 1 | NPTXR | 0.57 | 0.49 | 0.65 | NPTXR | 1.79E-01 | 1.5 (0.8-2.7) |
| 1 | FOLR1 | 0.55 | 0.44 | 0.66 | FOLR1 | 3.90E-01 | 1.5 (0.8-2.7) |
| 1 | CCL13 | 0.55 | 0.47 | 0.64 | CCL13 | 7.78E-01 | 1.2 (0.7-2.3) |
| 1 | PGF | 0.55 | 0.47 | 0.64 | PGF | 1.24E-01 | 1.5 (0.8-2.6) |
| 1 | IL15 | 0.55 | 0.47 | 0.63 | IL15 | 9.72E-01 | 1.1 (0.6-2) |
| 1 | PDGFRB | 0.55 | 0.47 | 0.63 | PDGFRB | 6.36E-01 | 1.1 (0.6-2.1) |
| 1 | VCAM1 | 0.53 | 0.45 | 0.64 | VCAM1 | 4.45E-01 | 1.5 (0.8-2.7) |
| 1 | AGRN | 0.53 | 0.46 | 0.62 | AGRN | 4.35E-01 | 1.3 (0.7-2.3) |
| 1 | UBB | 0.53 | 0.46 | 0.59 | UBB | 8.15E-01 | 0.9 (0.5-1.6) |
| 1 | IL13 | 0.53 | 0.46 | 0.60 | IL13 | 8.61E-01 | 1.1 (0.6-2) |
| 1 | IL16 | 0.53 | 0.44 | 0.62 | IL16 | 3.49E-01 | 1.6 (0.8-2.9) |

**Legend:** The tables present the area under the curve (AUC) with 95% confidence intervals (CI) for each NULISA CAF and plasma biomarker or biomarker combinations, together with the corresponding logistic regression coefficients. Prognostic value was further evaluated using Kaplan–Meier survival analysis with log-rank tests. Hazard ratios (HR) with 95% CI compare the 1st vs. 3rd tercile of the biomarker-derived score. The column *Num* indicates how many biomarkers are included in the model. The tables are sorted in AUC decreasing values.
